# DNA methylation as proxy of genetic, prenatal and perinatal psychiatric risk factors

**DOI:** 10.64898/2026.07.22.26358558

**Authors:** Elena Isaevska, Rosa H. Mulder, Isabel K. Schuurmans, Nicole Creasey, Janine F. Felix, Jean-Baptiste Pingault, Neeltje van Haren, Charlotte Cecil, Alexander Neumann

## Abstract

**Background:** Cord blood DNA methylation profile scores (MPSs) based on genetic and pre-/perinatal risk factors for neurodevelopmental conditions (NDCs) may capture downstream biological effects and help understand how combined exposure signals contribute to NDC risk.

**Methods:** Using data from two longitudinal birth cohorts, Generation R (N_train_ = 1856, N_test_ = 476) and ALSPAC (N_validation_= 832), we developed cord blood MPSs based on genetic and pre-/perinatal NDC risk factors. We assessed individual and combined predictive performance of risk factors and MPSs for eight childhood psychiatric outcomes (four broad, four specific), measured between ages 5 and 14 years. We also evaluated if the MPSs could be combined into a composite “transmission load” MPS.

**Results:** We validated four novel MPSs: maternal age, birthweight, and genetic liability for ADHD and schizophrenia (r range = 0.08 to 0.29) and included two previously validated MPSs: maternal smoking and gestational age (r range = 0.42 to 0.63). Jointly modeling the six MPSs with their corresponding risk factors explained on average 3.3% of variance in outcomes, higher than that explained by risk factors (1.8%) or MPSs alone (1.6%), indicating complementary sources of risk. The “transmission load” MPS did not replicate due to heterogeneous contributions of the predictors across cohorts.

**Conclusions:** The four novel MPSs based on genetic and pre-/perinatal risk factors can serve as valuable tools for future research. Integrating genetic and prenatal risk factors with DNA methylation at birth can provide insights into their individual and joint contributions to early psychiatric risk and may improve prediction.

## Introduction

Genetic, and pre-/perinatal risk factors are important contributors to neurodevelopmental (NDCs) and psychiatric conditions, but their downstream biological effects at birth remain difficult to characterize, limiting early risk assessment. Improving our ability to capture psychiatric risk as early as birth is important given their early and long-term impact on cognitive, behavioral and socioeconomic outcomes.^1^ Integrating risk factors across domains and diagnoses (given their transdiagnostic nature) may provide a more comprehensive representation of the “transmission load” at birth – the cumulative genetic, prenatal and perinatal factors involved in the intergenerational transmission of psychiatric risk.^2–4^

Previous attempts to integrate genetic and environmental exposures into risk assessment tools for neurodevelopmental and psychiatric disorders^5–7^ have been disorder-specific and rely on measuring multiple predictors. These can be difficult to obtain and subject to reporting biases, including recall bias and socially desirable responding. DNA methylation (DNAm) may help address some of these limitations as an epigenetic mechanism that regulates gene activity in response to both genetic and environmental factors^8^, thereby capturing their downstream biological effects at a molecular level.

DNAm scores, commonly referred to as Methylation Profile Scores (MPSs), aggregate information from many DNAm sites into a single score per individual, similar to polygenic scores (PGSs). Unlike PGS, MPSs are time-and tissue-specific, reflecting dynamic biological responses to exposures, making them useful for both past exposure assessment and future disease risk prediction.^9,10^ MPSs could offer advantages over assessing individual risk factors by providing continuous, objective measurements, improving retrospective assessment of long-term exposures and capturing the biological effect on the individual, not just their presence.^11,12^

DNAm at birth is associated with genetic liability to neurodevelopmental and psychiatric contitions^13^, such as ADHD, autism and schizophrenia, as well as a range of pre-/perinatal factors, such as maternal characteristics, pregnancy complications and birth outcomes^14–16^. This makes cord blood DNAm an attractive source for exposure assessment via MPSs, due to its temporal proximity to *in-utero* exposures and lack of postnatal influences. In addition, as only a single blood measurement is required to calculate multiple MPSs, it may be more cost-effective than assessing multiple exposures directly, that also require harmonization across cohorts.

Cord blood DNAm can also be used for prospective prediction, as it is measured before symptom onset. However, most MPSs are based on adult populations and single diagnoses, despite potential transdiagnostic effects. While there are notable examples of MPSs developed in newborns (gestational age^17,18^ and maternal smoking in pregnancy^19–21)^, validated pediatric MPSs for other genetic, pre-/perinatal exposures remain scarce.^22,23^ This further restricts our ability to evaluate how multiple exposure-related biological signals contribute to early psychiatric risk.

We aim to address these gaps by (1) constructing cord blood MPSs based on genetic, pre-/perinatal risk factors implicated in psychiatric risk and evaluating their performance in internal test and external validation datasets; (2) assessing their individual and combined predictive value for broad domains of childhood psychiatric problems: total, internalizing, externalizing and attention/hyperactivity problems, capturing underlying psychiatric liability across diagnoses, and specific domains: ADHD, autism and schizophrenia, while disentangling the relative contributions of MPSs and measured risk factors, as well as their integration for outcome prediction. As a final step (3) we combined these MPSs into a single composite score based on total childhood psychiatric problems, a proxy of “transmission load” risk, and evaluated its performance.

## Methods and Materials

The overall study design is summarized in **Figure 1**.

**Figure 1.**
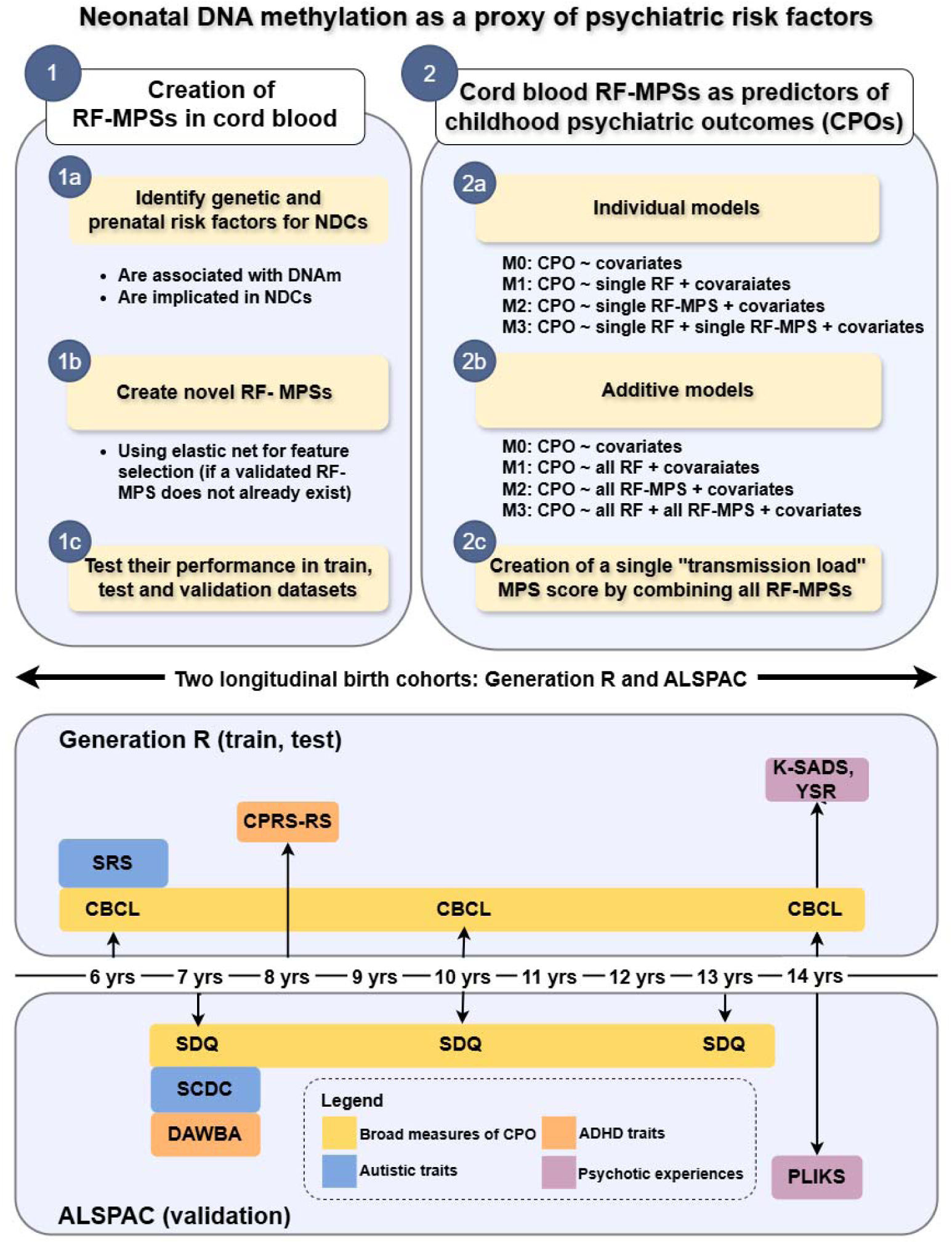
Outline of the analysis plan Abbreviations: RF: Risk Factor, MPS: Methylation Profile Scores, NDCs: Neurodevelopmental conditions; ADH: Attention Deficit Hyperactivity Disorder; CBCL: Child Behavior Checklist; SRS: Social Responsiveness Scale; CPRS-RS: Conners’ Parent Rating Scale-Revised Short Form; K-SADS: Kiddie-Schedule for Affective Disorders and Schizophrenia; YSR: Youth Self-Report; SDQ: Strengths and Difficulties Questionnaire; SCDC: Social Communication Disorder Checklist; DAWBA: Development and Well-Being Assessment; PLIKS: Psychosis-Like Experience

### Study population

The study includes data from two population-based birth cohorts: the Generation R Study^24–26^ (GenR) and the Avon Longitudinal Study of Parents and Children (ALSPAC). The MPSs were trained and internally tested in GenR and externally validated in ALSPAC.^27,28^ Briefly, GenR recruited 9778 pregnant women with delivery date between April 2002 and January 2006, residing in Rotterdam, the Netherlands. Pregnant women resident in Avon, UK with expected dates of delivery between 1st April 1991 and 31st December 1992 were invited to take part in the ALSPAC study. Both cohorts followed children and parents across multiple research waves that included questionnaires, anthropological measurements, genetic and epigenetic data. In the current study, we included children with available genetic and cord blood DNAm data from both cohorts. Only one child per family was included, specifically, the sibling with less missing data, or if equal, one sibling was kept at random. Full study details and flow chart describing the selection of study participants is available in **Supplementary Methods.**

### Identification of relevant genetic, prenatal and perinatal risk factors

We focused on ADHD, autism and schizophrenia, three psychiatric disorders with evidence of association with DNAm patterns at birth.^13^ We conducted two literature searches: one to identify pre-/perinatal factors linked to DNAm, and another to further narrow the search to transdiagnostic risk factors for the three conditions. We identified 11 genetic and perinatal risk factors that included: ADHD PGS, autism PGS, schizophrenia PGS, maternal pre-pregnancy BMI, maternal age, birthweight, hypertensive disorders in pregnancy, maternal diabetes, cesarean section, gestational age and maternal smoking. The literature search strategies and findings are provided in the **Supplementary Methods** and **Supplementary Tables S1-S2.**

### Assessment of genetic liability, prenatal and perinatal risk factors

Genotypes for PGS construction were derived from SNP arrays: HumanHap 610K and HumanHap 660W Quad chips in GenR, and the Illumina HumanHap550 quad chip in ALSPAC. The PGSs for ADHD, autism and schizophrenia were calculated using PRS-ice2^29^ and summary statistics from recent meta-analyses (Demontis et al., 2023; Grove et al., 2019; Trubetskoy et al., 2022). In both cohorts, information on all prenatal and perinatal risk factors was obtained through questionnaires, anthropometric measurements, or data from midwife and obstetric records. Further details on genotyping, quality control, imputation, PGS calculation using PRSice-2^13^, and the definition and measurement of risk factors is available in **Supplementary Methods**.

### Measurement of DNA methylation

DNAm was measured in cord blood in both cohorts. In GenR (train and test datasets) DNAm was measured using: (1) the Illumina Infinium HumanMethylation450 BeadChip (Illumina 450K), N_GenR-450K_ = 1393 or (2) the MethylationEPIC v1.0 BeadChip (Illumina EPIC), N_GenR-EPIC_ = 1115. Samples from both platforms were randomly allocated to the train and test datasets, such that each dataset contained a mixture of both 450K and EPIC arrays. In ALSPAC (the external validation dataset), DNAm was measured using the Illumina 450K array, N_ALSPAC_ = 913. In both cohorts, DNAm beta values were used. Detailed description of DNAm measurement in both cohorts, sample sizes, quality control and filtering steps, as well as normalization methods is provided in **Supplementary Methods.** Cross reactive probes were additionally removed using the *maxprobes* R-package^30^, while SNP-proximal probes were retained to preserve potentially predictive genetic signal. Only CpGs present both on EPIC and 450K arrays were included.

### Childhood psychiatric outcomes

To examine the prediction of traits capturing psychiatric liability across diagnoses, we included broad domains of childhood psychiatric problems, including total, internalizing, externalizing and attention/hyperactivity problems, measured in early childhood (6-8 years), late childhood (10 years) and early adolescence (13-14 years). To evaluate the prediction of specific phenotypic traits we included single-timepoint measures of ADHD traits, autism traits and psychotic symptoms, assessed at ages relevant for symptoms onset, **Figure 1**. More details on cohort specific questionnaires and outcome definitions are provided in **Supplementary Methods.**

### Covariates

Covariates included sex, maternal education, genetic ancestry (first four genomic principal components; PCs), child age at assessment of psychiatric problems, sample plate and six cell types estimated from DNAm values (CD8 T cells, CD4 T cells, natural killer cells, B cells, monocytes, and nucleated red blood cells; granulocytes were excluded to avoid multicollinearity). Further details on covariates are provided in **Supplementary Methods.**

### Missing data

Complete cases were used to train the novel MPS because missingness was below 10% for each risk factor and methods for handling missing data using regularized regression are less established. For all other analyses, missing data were imputed separately within each dataset (to avoid data leakage) using Multiple Imputation by Chained Equations (MICE) 100 imputation and 30 iterations, including all risk factors, outcomes and covariates. All reported results were pooled across imputations.

## Statistical analysis

### Step 1. Creating cord blood MPSs based on genetic, prenatal and perinatal risk factors

For risk factors without pre-existing validated MPS, we created novel risk-factor MPSs (RF-MPSs) using elastic-net regression implemented in the *glmnet* R-package. CpG-level summary statistics from epigenome-wide meta-analyses matching each risk factor were used for CpG pre-selection (nominal p-value <0.05), **Supplementary Table S3**. Elastic-net combines lasso and ridge penalties, which helps reduce overfitting and can handle correlated predictors, which is common when many CpGs are included. The mixing parameter (alpha) was fixed to 0.5 and the lambda parameter was selected through 10-fold cross-validation. CpGs and their weights were used to calculate RF-MPSs in the train, test and validation datasets. For risk factors with existing validated MPSs, RF-MPSs were calculated using previously published CpGs and weights. Performance was assessed through Mean Squared Error (MSE), Mean Absolute Error (MAE), correlations, covariate-adjusted partial correlations, and variance explained beyond the baseline covariates. The latter was quantified using partial R^2^, calculated as (R^2^_full_model_ - R^2^_baseline_model_)/(1 − R^2^_baseline_model_)^31^, from baseline (risk factor ∼ covariates) and a full (risk factor ∼ MPS + covariates) model.^31^

Only validated RF-MPSs, defined as those showing partial correlation p-value <0.05 in both the test and validation datasets, were included in the next steps that include prediction of childhood psychiatric outcome and construction of composite MPS.

We additionally performed functional analyses of the CpGs included in each validated RF-MPSs (novel or pre-existing) including: (1) pathway enrichment (GOmeth function, *missMethyl* R-package), (2) trait enrichment^32,33^ (3) enrichment analyses for CpGs at or near SNPs, methylation quantitative trait loci (mQTLs)^34^, CoRSIVs (“correlated regions of systemic interindividual variation”, known to have a strong genetic influence, high inter-individual variability and temporal stability)^35, 36^ and in imprint control regions^37^.

### Step 2. Individual and combined predictive utility of cord blood RF-MPSs for child psychiatric outcomes

To evaluate the predictive utility of RF-MPSs (and their contribution relative to and in combination with corresponding risk factors) we ran two sets of models: individual models (one RF-MPS and/or one risk factor) and combined models (all RF-MPSs and/or risk factors). Both model types were fitted in three ways: risk factors only (M1), RF-MPSs only (M2) and both (M3), **Figure 1**. Repeated childhood outcomes at three time points were analyzed using linear mixed models, whereas single time-point outcomes were analyzed using linear regression. Linear mixed models included a random intercept on participant level and were fit with REML as implemented in *lme4*. Partial R^2^ was calculated for each analysis using the pooled R^2^ values (using Fisher z-transformation in linear models and averaged across imputations for linear mixed models). For imputed data, nested model comparisons (comparing the fit of the full model M3 vs the simpler model M1) used D1-statistic as implemented in *mice* (linear models) and likelihood ratio test with *mitml* (linear mixed models), complementary to anova in non-imputed models.

We additionally performed dominance analysis implemented in *dominanceanalysis* which disentangles the average contribution of each predictor to R^2^ across all possible subset models, while holding covariates constant. All variables were z-score standardized prior to analysis to ensure comparability.

### Step 3. Creating composite score, based on MPSs and predictive of childhood psychiatric problems

Next, we trained a new integrative score using all validated (pre-existing or novel) RF-MPSs to capture overall “transmission load” (TL score) predictive of childhood psychiatric problems, as measured by the total problems score. To identify the optimal combination of RF-MPS we applied an elastic-net regression in the train dataset using all RF-MPS as predictors of total child problems. The resulting TL score is intended to reflect the combined load of genetic, prenatal and perinatal risk factors using minimal number of predictors, derived solely on DNAm at birth. We then evaluated its performance in the test and validation datasets.

## Results

A total of 3164 participants were included in the study, 1856 in the train dataset, 476 in the test and 832 in the validation dataset. The three datasets were similar in the distribution of the risk factors, see **Table 1, Supplementary Figure S1-S4**.

**Table 1.**
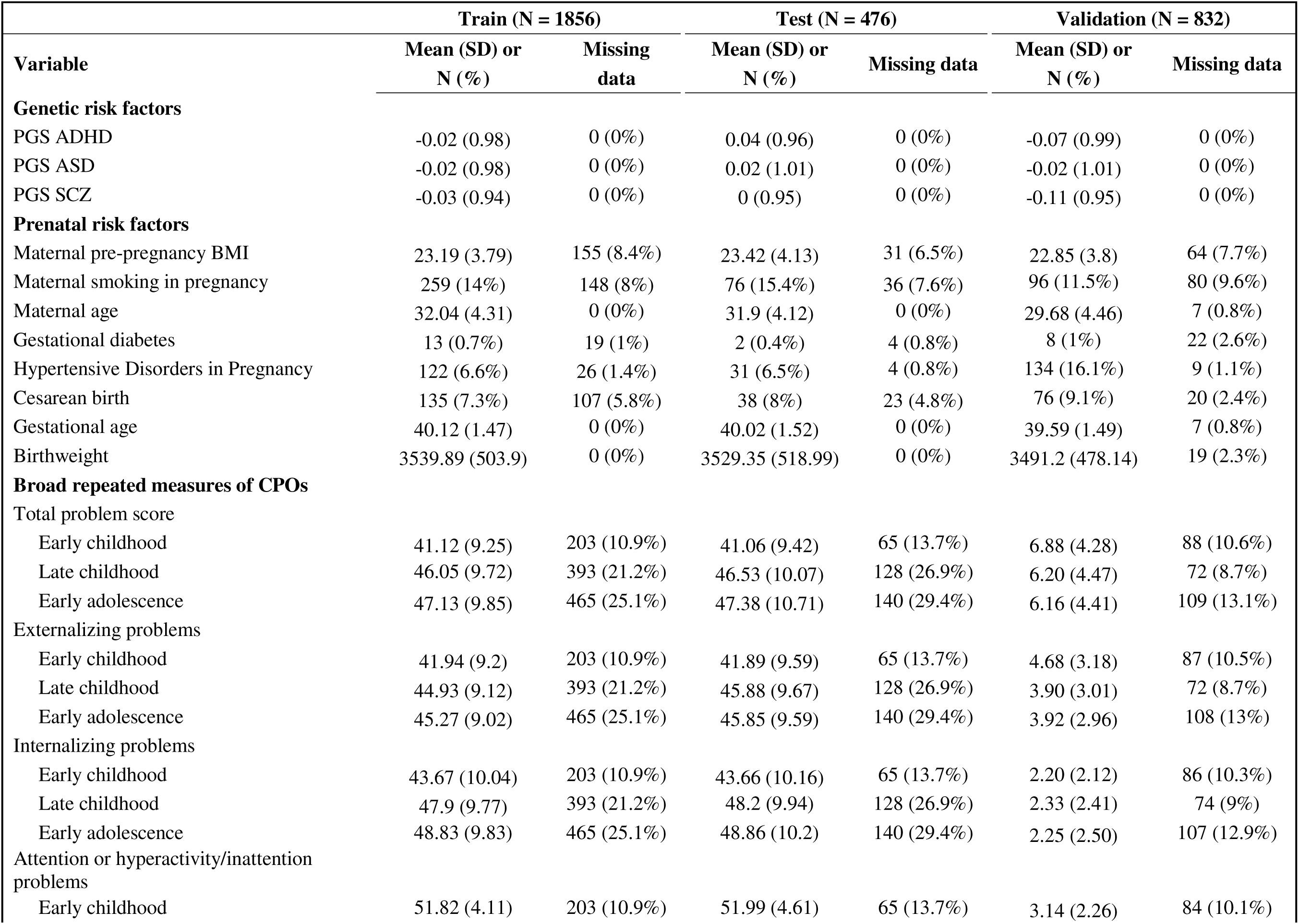
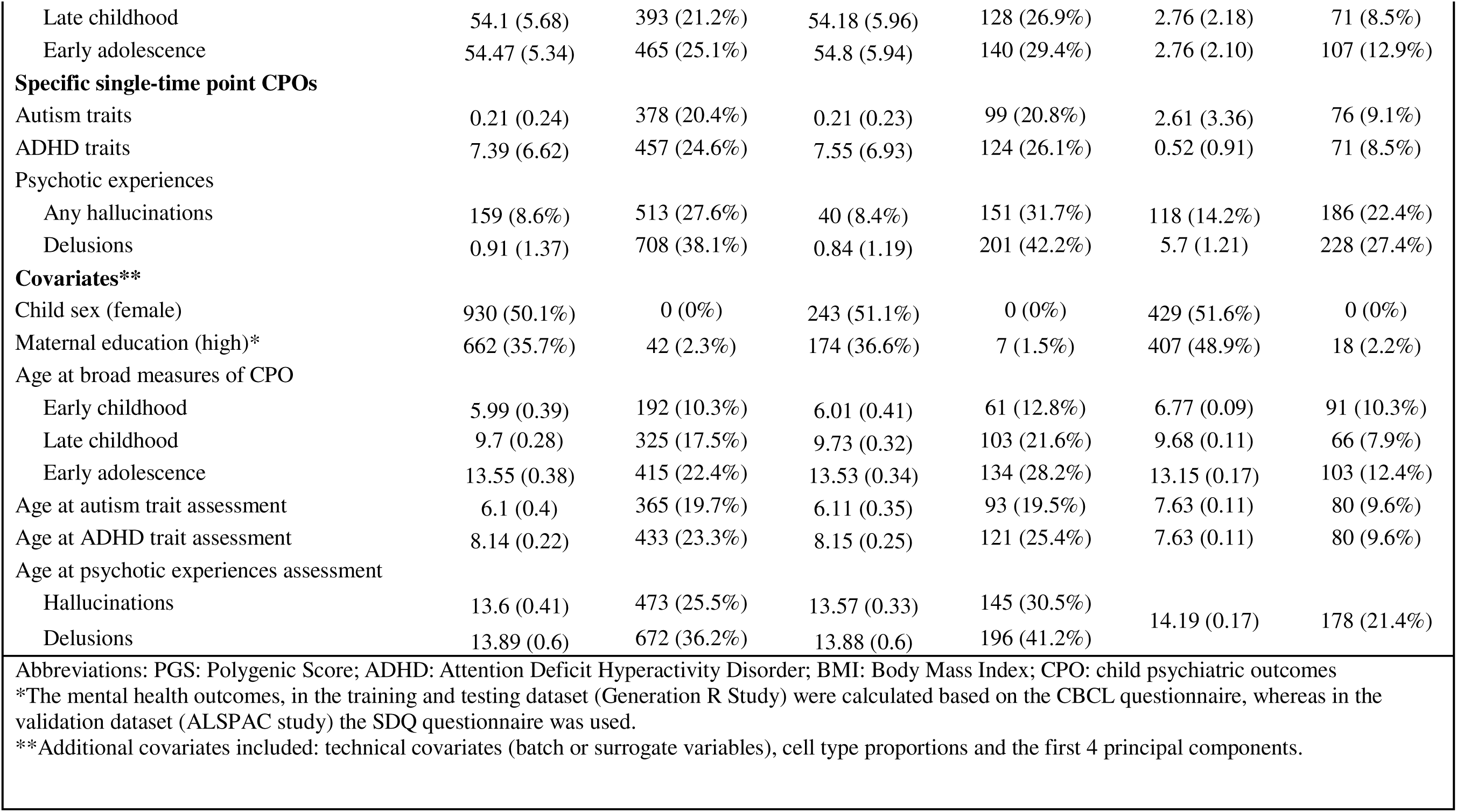
Population characteristics of the three datasets included in the study.

| Variable | Train (N = 1856) |  | Test (N = 476) |  | Validation (N = 832) |  |
| --- | --- | --- | --- | --- | --- | --- |
|  | Mean (SD) or<br>N (%) | Missing<br>data | Mean (SD) or<br>N (%) | Missing data | Mean (SD) or<br>N (%) | Missing data |
| **Genetic risk factors** |  |  |  |  |  |  |
| PGS ADHD | -0.02 (0.98) | 0 (0%) | 0.04 (0.96) | 0 (0%) | -0.07 (0.99) | 0 (0%) |
| PGS ASD | -0.02 (0.98) | 0 (0%) | 0.02 (1.01) | 0 (0%) | -0.02 (1.01) | 0 (0%) |
| PGS SCZ | -0.03 (0.94) | 0 (0%) | 0 (0.95) | 0 (0%) | -0.11 (0.95) | 0 (0%) |
| **Prenatal risk factors** |  |  |  |  |  |  |
| Maternal pre-pregnancy BMI | 23.19 (3.79) | 155 (8.4%) | 23.42 (4.13) | 31 (6.5%) | 22.85 (3.8) | 64 (7.7%) |
| Maternal smoking in pregnancy | 259 (14%) | 148 (8%) | 76 (15.4%) | 36 (7.6%) | 96 (11.5%) | 80 (9.6%) |
| Maternal age | 32.04 (4.31) | 0 (0%) | 31.9 (4.12) | 0 (0%) | 29.68 (4.46) | 7 (0.8%) |
| Gestational diabetes | 13 (0.7%) | 19 (1%) | 2 (0.4%) | 4 (0.8%) | 8 (1%) | 22 (2.6%) |
| Hypertensive Disorders in Pregnancy | 122 (6.6%) | 26 (1.4%) | 31 (6.5%) | 4 (0.8%) | 134 (16.1%) | 9 (1.1%) |
| Cesarean birth | 135 (7.3%) | 107 (5.8%) | 38 (8%) | 23 (4.8%) | 76 (9.1%) | 20 (2.4%) |
| Gestational age | 40.12 (1.47) | 0 (0%) | 40.02 (1.52) | 0 (0%) | 39.59 (1.49) | 7 (0.8%) |
| Birthweight | 3539.89 (503.9) | 0 (0%) | 3529.35 (518.99) | 0 (0%) | 3491.2 (478.14) | 19 (2.3%) |
| **Broad repeated measures of CPOs** |  |  |  |  |  |  |
| Total problem score |  |  |  |  |  |  |
| Early childhood | 41.12 (9.25) | 203 (10.9%) | 41.06 (9.42) | 65 (13.7%) | 6.88 (4.28) | 88 (10.6%) |
| Late childhood | 46.05 (9.72) | 393 (21.2%) | 46.53 (10.07) | 128 (26.9%) | 6.20 (4.47) | 72 (8.7%) |
| Early adolescence | 47.13 (9.85) | 465 (25.1%) | 47.38 (10.71) | 140 (29.4%) | 6.16 (4.41) | 109 (13.1%) |
| Externalizing problems |  |  |  |  |  |  |
| Early childhood | 41.94 (9.2) | 203 (10.9%) | 41.89 (9.59) | 65 (13.7%) | 4.68 (3.18) | 87 (10.5%) |
| Late childhood | 44.93 (9.12) | 393 (21.2%) | 45.88 (9.67) | 128 (26.9%) | 3.90 (3.01) | 72 (8.7%) |
| Early adolescence | 45.27 (9.02) | 465 (25.1%) | 45.85 (9.59) | 140 (29.4%) | 3.92 (2.96) | 108 (13%) |
| Internalizing problems |  |  |  |  |  |  |
| Early childhood | 43.67 (10.04) | 203 (10.9%) | 43.66 (10.16) | 65 (13.7%) | 2.20 (2.12) | 86 (10.3%) |
| Late childhood | 47.9 (9.77) | 393 (21.2%) | 48.2 (9.94) | 128 (26.9%) | 2.33 (2.41) | 74 (9%) |
| Early adolescence | 48.83 (9.83) | 465 (25.1%) | 48.86 (10.2) | 140 (29.4%) | 2.25 (2.50) | 107 (12.9%) |
| Attention or hyperactivity/inattention problems |  |  |  |  |  |  |
| Early childhood | 51.82 (4.11) | 203 (10.9%) | 51.99 (4.61) | 65 (13.7%) | 3.14 (2.26) | 84 (10.1%) |
| Late childhood | 54.1 (5.68) | 393 (21.2%) | 54.18 (5.96) | 128 (26.9%) | 2.76 (2.18) | 71 (8.5%) |
| Early adolescence | 54.47 (5.34) | 465 (25.1%) | 54.8 (5.94) | 140 (29.4%) | 2.76 (2.10) | 107 (12.9%) |
| Specific single-time point CPOs |  |  |  |  |  |  |
| Autism traits | 0.21 (0.24) | 378 (20.4%) | 0.21 (0.23) | 99 (20.8%) | 2.61 (3.36) | 76 (9.1%) |
| ADHD traits | 7.39 (6.62) | 457 (24.6%) | 7.55 (6.93) | 124 (26.1%) | 0.52 (0.91) | 71 (8.5%) |
| Psychotic experiences |  |  |  |  |  |  |
| Any hallucinations | 159 (8.6%) | 513 (27.6%) | 40 (8.4%) | 151 (31.7%) | 118 (14.2%) | 186 (22.4%) |
| Delusions | 0.91 (1.37) | 708 (38.1%) | 0.84 (1.19) | 201 (42.2%) | 5.7 (1.21) | 228 (27.4%) |
| Covariates** |  |  |  |  |  |  |
| Child sex (female) | 930 (50.1%) | 0 (0%) | 243 (51.1%) | 0 (0%) | 429 (51.6%) | 0 (0%) |
| Maternal education (high)* | 662 (35.7%) | 42 (2.3%) | 174 (36.6%) | 7 (1.5%) | 407 (48.9%) | 18 (2.2%) |
| Age at broad measures of CPO |  |  |  |  |  |  |
| Early childhood | 5.99 (0.39) | 192 (10.3%) | 6.01 (0.41) | 61 (12.8%) | 6.77 (0.09) | 91 (10.3%) |
| Late childhood | 9.7 (0.28) | 325 (17.5%) | 9.73 (0.32) | 103 (21.6%) | 9.68 (0.11) | 66 (7.9%) |
| Early adolescence | 13.55 (0.38) | 415 (22.4%) | 13.53 (0.34) | 134 (28.2%) | 13.15 (0.17) | 103 (12.4%) |
| Age at autism trait assessment | 6.1 (0.4) | 365 (19.7%) | 6.11 (0.35) | 93 (19.5%) | 7.63 (0.11) | 80 (9.6%) |
| Age at ADHD trait assessment | 8.14 (0.22) | 433 (23.3%) | 8.15 (0.25) | 121 (25.4%) | 7.63 (0.11) | 80 (9.6%) |
| Age at psychotic experiences assessment |  |  |  |  |  |  |
| Hallucinations | 13.6 (0.41) | 473 (25.5%) | 13.57 (0.33) | 145 (30.5%) | 14.19 (0.17) | 178 (21.4%) |
| Delusions | 13.89 (0.6) | 672 (36.2%) | 13.88 (0.6) | 196 (41.2%) |  |  |
| Abbreviations: PGS: Polygenic Score; ADHD: Attention Deficit Hyperactivity Disorder; BMI: Body Mass Index; CPO: child psychiatric outcomes |  |  |  |  |  |  |
| *The mental health outcomes, in the training and testing dataset (Generation R Study) were calculated based on the CBCL questionnaire, whereas in the validation dataset (ALSPAC study) the SDQ questionnaire was used. |  |  |  |  |  |  |
| **Additional covariates included: technical covariates (batch or surrogate variables), cell type proportions and the first 4 principal components. |  |  |  |  |  |  |

### Cord blood MPSs as surrogates of genetic and prenatal risk factors (RF-MPSs)

We created nine novel RF-MPSs in the train dataset: genetic liability ADHD, autism and schizophrenia, maternal age, maternal pre-pregnancy BMI, maternal age, birthweight, hypertensive disorders in pregnancy, gestational diabetes and cesarean birth. We also calculated two previously validated RF-MPSs for maternal smoking in pregnancy and gestational age, **Table 2**. Of the nine novel RF-MPSs, four were fully validated, showing nominally significant correlations with their respective risk factors independent of covariates, in both the test and the validation dataset (partial correlation p value < 0.05): ADHD PGS, schizophrenia PGS, maternal age and birthweight. The numbers included in the validated RF-MPSs ranged from 2346 for birthweight to 99 for schizophrenia PGS. Maternal pre-pregnancy BMI and cesarean section showed initial correlations that attenuated after covariate adjustment and therefore did not meet the criteria for full validation. The RF-MPSs for autism PGS, gestational diabetes and hypertensive disorders in pregnancy were not validated. The CpGs and the corresponding weights, as well as the performance metrics for all RF-MPSs can be found in **Supplementary Tables S4-S5**, **Supplementary Figure 5-8.**

**Table 2.**
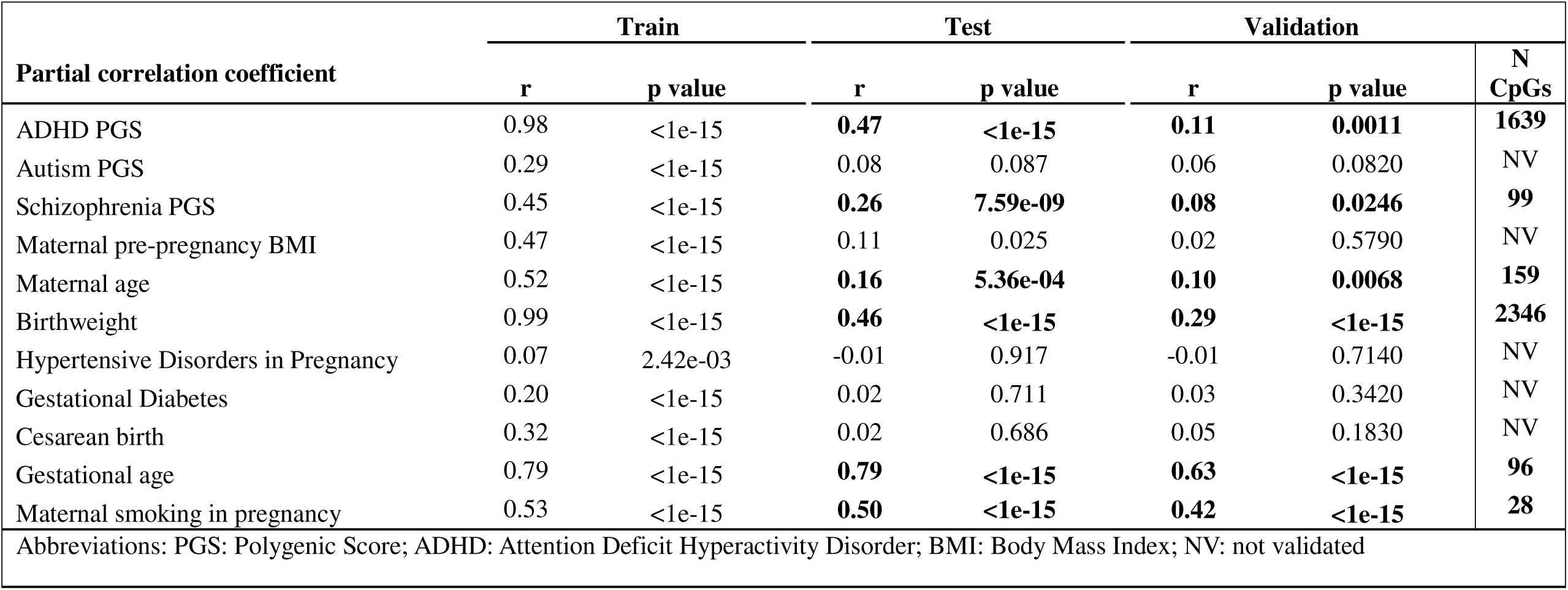
Partial correlation coefficients (adjusted for baseline covariates) between the genetic and prenatal risk factors and their respective MPSs in the test and training dataset. Imputed dataset was used to derive the coefficients. Gestational age and maternal smoking MPSs were created and validated in previous studies. RF-MPSs included in subsequent analyses are shown in bold.

| Partial correlation coefficient | Train |  | Test |  | Validation |  | N<br>CpGs |
| --- | --- | --- | --- | --- | --- | --- | --- |
|  | r | p value | r | p value | r | p value |  |
| ADHD PGS | 0.98 | <1e-15 | **0.47** | **<1e-15** | **0.11** | **0.0011** | **1639** |
| Autism PGS | 0.29 | <1e-15 | 0.08 | 0.087 | 0.06 | 0.0820 | NV |
| Schizophrenia PGS | 0.45 | <1e-15 | **0.26** | **7.59e-09** | **0.08** | **0.0246** | **99** |
| Maternal pre-pregnancy BMI | 0.47 | <1e-15 | 0.11 | 0.025 | 0.02 | 0.5790 | NV |
| Maternal age | 0.52 | <1e-15 | **0.16** | **5.36e-04** | **0.10** | **0.0068** | **159** |
| Birthweight | 0.99 | <1e-15 | **0.46** | **<1e-15** | **0.29** | **<1e-15** | **2346** |
| Hypertensive Disorders in Pregnancy | 0.07 | 2.42e-03 | -0.01 | 0.917 | -0.01 | 0.7140 | NV |
| Gestational Diabetes | 0.20 | <1e-15 | 0.02 | 0.711 | 0.03 | 0.3420 | NV |
| Cesarean birth | 0.32 | <1e-15 | 0.02 | 0.686 | 0.05 | 0.1830 | NV |
| Gestational age | 0.79 | <1e-15 | **0.79** | **<1e-15** | **0.63** | **<1e-15** | **96** |
| Maternal smoking in pregnancy | 0.53 | <1e-15 | **0.50** | **<1e-15** | **0.42** | **<1e-15** | **28** |
| Abbreviations: PGS: Polygenic Score; ADHD: Attention Deficit Hyperactivity Disorder; BMI: Body Mass Index; NV: not validated |  |  |  |  |  |  |  |

In terms of partial correlations for the novel and validated RF-MPSs, coefficients in the test set ranged from r=0.16 (maternal age; p-value 5.3 x 10^-04^) to r=0.47 (ADHD PGS, p-value <1×10^-15^). In the external validation dataset, they were lower, ranging from r=0.08, (schizophrenia PGS, p-value 0.02) to r=0.29, (birthweight, p-value <1×10^-15^). The largest attenuation across datasets was observed for ADHD PGS (r = 0.98 in train, r = 0.47 in test, and r=0.11 in external validation). The previously validated RF-MPSs (maternal smoking and gestational age) showed comparable correlations with their corresponding phenotypes across GenR and ALSPAC, with r values ranging from 0.42 to 0.53 for maternal smoking and from 0.63 to 0.79 for gestational age, **Table 2**. The RF-MPS for birthweight correlated with the RF-MPS for gestational age (r=0.43 in test, and r=0.29 in validation dataset), similar to the correlation in risk factors (r=0.53 in test, r=0.39 in validation dataset), despite minimal overlap in CpGs, **Supplementary Table S5-8.** The RF-MPS for gestational age had the highest explained variance in the external validation dataset (39%), followed by maternal smoking (20%), birthweight (9%), and ADHD PGS, schizophrenia PGS and maternal age (1%), **Figure 2, Supplementary Table S10.**

**Figure 2.**
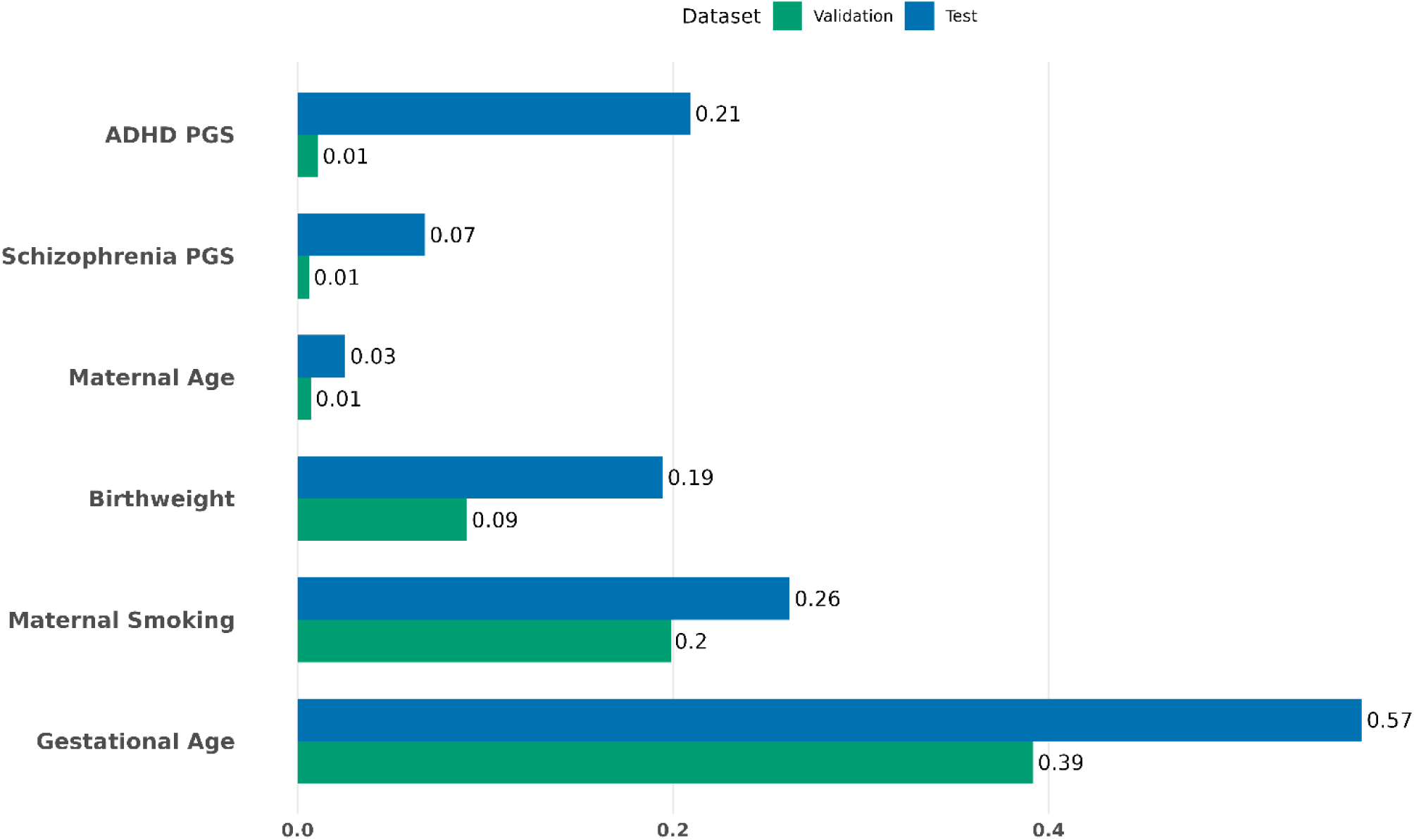
Variance in genetic and pre-/perinatal risk factors factors explained by their respective validated methylation profile scores in terms of partial R^2^.

The functional analysis of the CpGs included in the MPSs, did not reveal significantly enriched pathways, but enriched traits were identified, including prenatal exposures and NDCs. Additionally, the RF-MPSs showed enrichment in mQTLs, CoRSIVs and imprint control regions. Detailed results are reported in **Supplementary Table S7-S9.**

### Individual predictive utility of RF-MPSs for childhood psychiatric outcomes

In the “risk-factor only” model (M1), the average partial R^2^ across the datasets and outcomes was 0.33% (SD=0.40). The RF-MPS model (M2), showed a slightly lower average partial R^2^ of 0.27% (SD=0.42), **Figure 3**, **Supplementary Table S11**. Explained variance increased in to 0.56% (SD=0.58) when modeling the risk factor and its RF-MPSs together. Modeling risk factors together with their corresponding RF-MPS appeared to improve model fit; however, most improvements were not statistically significant, except the gestational age RF-MPS and externalizing problems in the test set (R^2^=1.66%, p=0.025). Notably, the RF-MPS for maternal smoking had one of the largest and most consistent associations with an autism score in both test (M2 R^2^=0.61%) and validation set (M2 R^2^=1.51%), but not reaching statistical significance in either.

**Figure 3.**
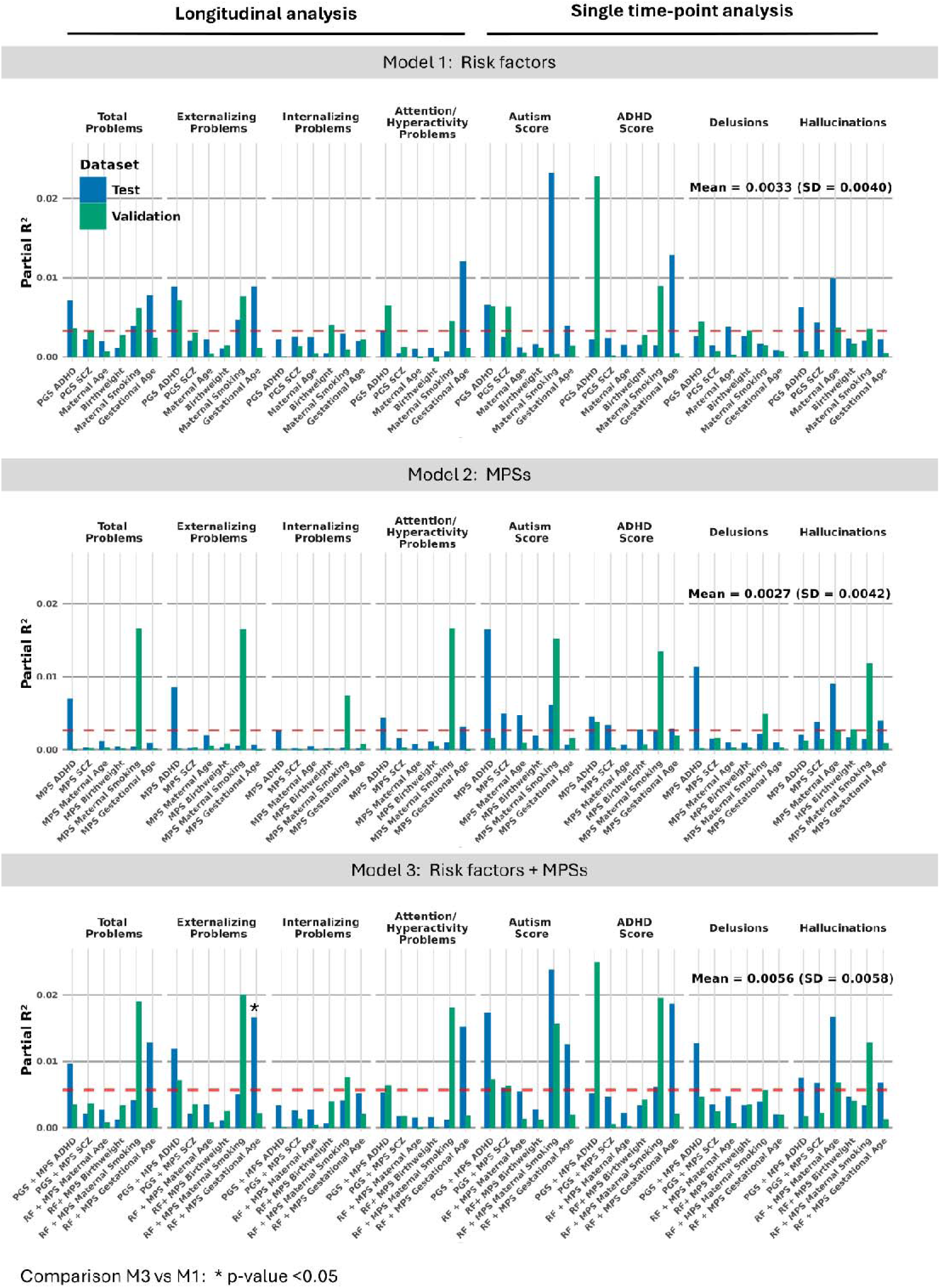
The figure shows the partial R^2^ from the individual models across datasets and three model types: model including single genetic or pre-/perinatal risk factor (Model 1), model including single RF-MPSs (Model 2), and model th t combines the risk factor with the corresponding RRF-MPS (Model 3).

### Additive predictive utility of RF-MPSs for childhood psychiatric outcomes

In the additive models, the results from the M1, M2 and M3 show similar patterns to the one from the individual models. The combination of all six RF-MPS shows a slightly lower partial R^2^ (mean 1.61%, SD 0.76) than the combination of all six risk factors (mean 1.81%, SD 0.79), but their combination produces the largest explained variance (mean 3.28%, SD 1.23). The highest partial R^2^ was seen for model M3 for autism score in the test dataset (partial R^2^=5.99%) and ADHD score in the validation dataset (partial R^2^=5.09%). Internalizing problems showed the smallest variance explained across all three models and two datasets, **Figure 4, Supplementary Table S12**. Although adding RF-MPSs showed a nearly additive effect, i.e. on average partial R^2^ for M3 were approximately the sum of M1 and M2 across outcomes, this improvement was not statistically significant.

**Figure 4.**
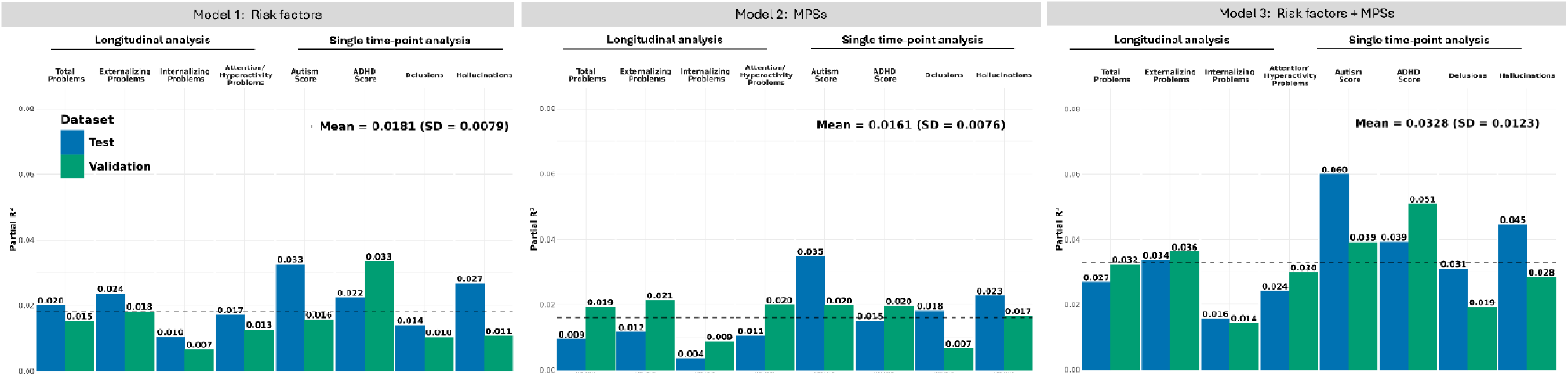
The figure shows the partial R^2^ from the combined models across two datasets (test and validation) and the three model types: model including all individual genetic and pre-/perinatal risk factors (Model 1), model including all individual RF-MPSs (Model 2), and model that combines both (Model 3).

The dominance analysis enabled examination of individual predictor contributions to explained variance. The contributions of predictors were heterogenous across datasets/cohorts, **Figure 5**. For example, in the test set, a major contributor to the variance in the total, externalizing and attention/hyperactivity problems were gestational age and RF-MPS for ADHD PGS, whereas in the validation dataset top contributors were ADHD PGS, maternal smoking and maternal smoking RF-MPS, **Supplementary Table S13**.

**Figure 5.**
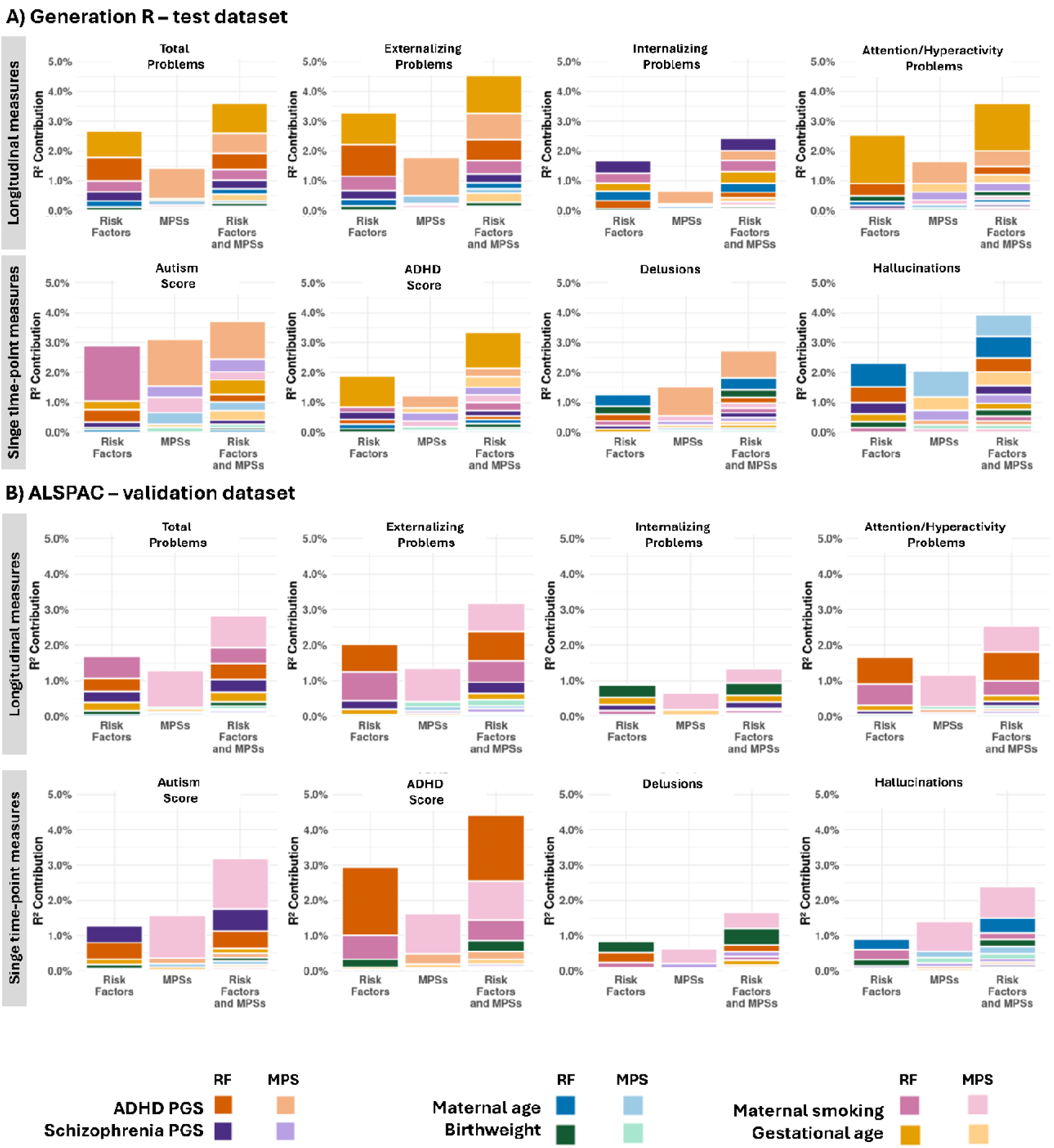
The results from the dominance analysis, showing the individual contribution of various risk factors and RF-MPS to the R^2^ (beyond the baseline covariates), are presented across three models: model including all individual genetic and pre-/perinatal risk factors (Model 1), model including all individual RF-MPSs (Model 2), and model that combines both (Model 3).

### Creating a composite transmission-load score (TL-score) that integrates all measures

We computed a composite transmission load score by combining the MPSs into a single score in the train dataset. We were able to replicate the signal in the internal test, but not in the external validation dataset. Coefficient estimates to calculate the TL-MPS are shown in **Supplementary Table S14**).

## Discussion

In this study, using data from two prospective population-based birth cohorts, we evaluated whether neonatal MPSs capture molecular signatures of genetic and pre-/perinatal risk factors and improve the early prediction of childhood psychiatric symptoms. We constructed and validated four novel neonatal RF-MPSs and found that, although the RF-MPSs contributed modestly to the explained variance, they captured signals distinct from the measured risk factors, as their combination nearly doubled the variance explained. Finally, we developed an integrative score TL-score based on RF-MPSs, which performed poorly in predicting total childhood psychiatric problems, likely due to the heterogeneity in the relative contribution of individual RF-MPSs across cohorts.

The development of these RF-MPSs represents an important step, as no RF-MPSs have previously been reported for maternal age or genetic liability to ADHD and schizophrenia^22,23^. Although a birthweight RF-MPS has been developed previously^38^, it lacked external validation and explained less variance (R^2^ = 0.14% in child buccal cells and 0.39% in adult whole blood) than our model (partial R^2^ = 8.82% in cord blood, validation dataset), although the differences in tissue type and measurement age may contribute. Notably, birthweight and gestational age RF-MPSs captured a substantial proportion of the DNAm – explained variance. DNAm has been estimated to explain 12.9% of birthweight variance (23.2% in GenR and 8.5% in ALSPAC) and 45.8% of gestational age variance (63.0% in GenR and 37.5% in ALSPAC).^39^ The variance explained by our birthweight RF-MPS (19.60% in GenR and 8.82% in ALSPAC) and gestational age RF-MPS (57.67% in GenR and 38.96% in ALSPAC) falls close to these ranges.

In contrast, we were unable to develop robust RF-MPSs for some risk factors, possibly due to overfitting to the GenR population and limiting generalizability across cohorts. Limited case numbers additionally hindered the development of robust RF-MPSs for gestational diabetes and hypertensive disorders in pregnancy. For maternal pre-pregnancy BMI and cesarean section, correlations attenuated after adjustment for baseline covariates, suggesting that these RF-MPSs may capture broader participant characteristics rather than a risk factor-specific signal. Interestingly, robust RF-MPS replication was not predicated on EWAS signal strength. For example, the EWAS on maternal pre-pregnancy BMI had more CpGs with nominal significance (94,009) than maternal age (25,879) or birthweight (50,875).

Next, we assessed whether validated RF-MPSs not only reflect pre-/perinatal exposures, but also predict child psychiatric problems, four of which were repeatedly measured. The variance explained by individual risk factors and RF-MPSs across all outcomes was modest (mean partial R^2^ <1%). However, selected risk factors with their corresponding RF-MPSs (eg. ADHD PGS, maternal smoking and gestational age) resulted in a notable increase in partial R^2^ compared to a risk-factor only model. While a lack of statistical significance for most risk factors limits firm conclusions, a replicable effect across both studies for maternal smoking RF-MPS and autistic scores is notable. Models combining all six risk factor and/or RF-MPSs, explained more variance than individual models, with combined M3 models showing the highest explained variance (mean partial R^2^=3.28%). Notably, averaged across outcomes, this increase in variance was a near perfect summation of the explained variance of a DNAm-based model (R^2^=1.61%) and RF only models (R^2^=1.81%). While the lack of statistical significance requires caution, the doubling of explained variance indicates potential for independent risk prediction of MPS- and risk factor-based predictors.

Although direct comparisons are limited, the magnitude of variance explained in our study is broadly comparable to previous work. In another study, PGS ADHD combined with an environmental risk variable score that included prenatal and postnatal variables, explained 3.19% and 3.51% of SDQ’s hyperactivity scales and CBCL attention scores, respectively. In our study, the combined risk factor and RF-MPS model explained 2.41% of CBCL’s attention score (GenR test set) and 2.98% of SDQ’s hyperactivity scale (ALSPAC validation dataset). For ADHD trait scores, the combined models explained 3.92% of ADHD scores in GenR test dataset and 5.09% in ALSPAC validation dataset.^40^ The complementary variance explained by risk factors and RF-MPS is consistent with adult studies on MPS for BMI, where variance explained was 7%, 8% and 14% for the MPS, PGS and a combined model, respectively.^41^

Finally, we aimed to create a single prediction score, or “transmission load” score, reflecting genetic and pre-/perinatal risk for total child psychiatric problems. The score did not show predictive performance in the external validation dataset. This likely reflects varying predictor contributions across cohorts, as demonstrated by the dominance analysis. This variability in contributions may be driven by sampling variability and population or methodological differences. As a result, it was difficult to derive a stable combination of RF-MPS weights that could generalize to the independent cohort. Another study also tried integrating genetic, epigenetic and metabolomic data for ADHD prediction using a different methodological framework, but failed to see a gain in explained variance in an external dataset.^42^

This study has several strengths, which include the integration of genetic and pre-/perinatal risk factors with DNAm, the prospective design and the comprehensive outcome assessments across childhood, and the use of an external validation cohort.^43^ The four newly developed RF-MPSs performed well across two cohorts from different countries and enrollment periods, array platforms and different normalization pipelines (quantile normalization in GenR and functional normalization in ALSPAC). Future studies should examine their performance and generalizability in other populations and newer platforms, such as Infinium MethylationEPIC v2.0.^44^ Further work is also needed to evaluate the role of RF-MPSs both as biological surrogates and complementary variables to measured risk factors.

The study also has some limitations. The RF-MPSs were developed in a European population, which may limit their generalizability to populations of other ancestries. All MPSs were developed and tested in cord blood and their applicability in other tissues and time points may be limited. GenR and ALSPAC contributed to the EWAS meta-analyses used for CpG preselection, but overfitting is unlikely since CpGs were only used for preselection, not weighting, and GenR’s or ALSPAC’s contribution (with the exception of gestational diabetes) never exceeded 15% of the total sample size, and furthermore GenR-EPIC was not included in the meta-analyses. Despite including a comprehensive set of childhood psychiatric outcome and risk factors, other relevant prenatal risk factors may have been missed because they have not been yet examined in relation to DNAm, or studies were underpowered to detect associations, and therefore did not meet our inclusion criteria. Incorporating postnatal risk factors and corresponding MPSs could further increase predictive utility.

By integrating genetic, pre-/perinatal risk factors and DNAm data, our study provides insights into early risk prediction of childhood psychiatric problems and provides a framework for combining multi-domain risk indicators at birth. We calculated six cord blood MPSs, four of which are novel additions to the developmental MPS atlas: genetic liability for ADHD, genetic liability for schizophrenia, birthweight and maternal age. The MPSs appear to explain unique variance in child psychiatric outcomes over and above the corresponding measured risk factors, but lack of statistical precision requires further replication. Integrating genetic and pre-/perinatal risk factors with DNAm at birth shows early promise for improving prediction and provides valuable insights into their individual contributions to early psychiatric risk, while also highlighting limitations in cross-cohort generalizability.

## Data Availability

Data from this study are available upon reasonable request to the director of the Generation R Study, subject to local, national and European rules and regulations. The Medical Ethical Committee of Erasmus MC, University Medical Center Rotterdam has approved the Generation R Study.

The informed consent obtained from ALSPAC (Avon Longitudinal Study of Parents and Children) participants does not allow the data to be made available through any third party maintained public repository. Supporting data are available from ALSPAC on request under the approved proposal number, B4611. Full instructions for applying for data access can be found here: http://www.bristol.ac.uk/alspac/researchers/access/. The ALSPAC study website contains details of all available data (http://www.bristol.ac.uk/alspac/researchers/our-data/).

## Acknowledgements

The project was funded by the European Union’s HorizonEurope Research and Innovation Programme (FAMILY, Grant Agreement No. 101057529). The views and opinions expressed are, however, those of the author(s) only and do not necessarily reflect those of the European Union or the European Health and Digital Executive Agency. Neither the European Union nor the granting authority can be held responsible for them. The work of CAMC is supported by the European Union’s HorizonEurope Research and Innovation Programme (HappyMums, Grant Agreement No. 101057390) and the European Research Council (TEMPO, Grant Agreement No. 101039672, also AN). This research was conducted while CAMC was a Hevolution/AFAR New Investigator Awardee in Aging Biology and Geroscience Research.

The general design of the Generation R Study is made possible by financial support from Erasmus MC, Erasmus University Rotterdam, the Netherlands Organization for Health Research and Development, and the Ministry of Health, Welfare and Sport. The EWAS data were funded by a grant from the Netherlands Genomics Initiative/Netherlands Organization for Scientific Research Netherlands Consortium for Healthy Aging (project No. 050-060-810), by funds from the Genetic Laboratory of the Department of Internal Medicine, Erasmus MC, and by a grant from the National Institute of Child and Human Development (Grant No. R01HD068437). The normalization of the Generation R EPIC data was supported by Helmholtz International Fellow Award HIFA-0174-RA-30/19

The UK Medical Research Council and Wellcome (Grant ref: MR/Z505924/1) and the University of Bristol provide core support for ALSPAC. A comprehensive list of grants funding is available on the ALSPAC website (http://www.bristol.ac.uk/alspac/external/documents/grant-acknowledgements.pdf). This publication is the work of the authors and Alexander Neumann and Elena Isaevska will serve as guarantors for the contents of this paper.

The Generation R Study is conducted by Erasmus MC University Medical Center Rotterdam, in close collaboration with the School of Law and Faculty of Social Sciences of the Erasmus University Rotterdam, the Municipal Health Service Rotterdam area, Rotterdam, the Rotterdam Homecare Foundation, Rotterdam and the Stichting Trombosedienst & Artsenlaboratorium Rijnmond, Rotterdam. We gratefully acknowledge the contribution of children and parents, general practitioners, hospitals, midwives, and pharmacies in Rotterdam. The generation and management of the DNA methylation data for the Generation R Study was executed by the Human Genotyping Facility of the Genetic Laboratory of the Department of Internal Medicine, Erasmus MC, the Netherlands. We thank Mr. Michael Verbiest, Ms. Mila Jhamai, Ms. Sarah Higgins, Mr. Marijn Verkerk, and Dr. Lisette Stolk for their help in creating the EWAS database. We thank Dr. A. Teumer for his work on the quality control and normalization scripts.

We are extremely grateful to all the families who took part in this study, the midwives for their help in recruiting them, and the whole ALSPAC team, which includes data collection staff, data and administrations staff, technical managers and the technical staff with the Bristol Bioresource Laboratory, based within the University of Bristol.

