## Supplementary Methods for "DNA methylation as proxy of genetic, prenatal and perinatal psychiatric risk factors"

### Study Population

#### The Generation R Study

The Generation R Study (GenR) is a population-based prospective cohort from fetal life onwards. In short, pregnant women were eligible for study participation if they were residents of the study area (Rotterdam, the Netherlands), and if they had a delivery date between April 2002 and January 2006.[1–3] A total of 9,778 mothers were enrolled, gave birth to 9749 live born children. These mothers, their children and partners took part in several research waves.

At the age of 6 years (early school age), we invited all 9278 children from the original cohort of 9749 children to participate in follow-up studies. This invitation was independent of their home address and participation in the preschool period. In total, 8305 children (90% of those who were invited (n = 9278) and 85% of the original cohort (n = 9749)) still participated in the study at this age, of whom 6690 visited the research center at a median age of 6.0 years. For the follow-up phase at the age of 10 years (mid childhood period) 730 children of the 9278 could not be invited. In total, 7393 children (86% of those who were invited (n = 8548) and 76% of the original cohort (n = 9749)) participated in the study in mid childhood, of whom 5862 visited the research center at a median age of 9.7 years. Of the 8548 children invited in the mid childhood period, 456 had withdrawn and 124 children were lost to follow-up during this period, leaving 7968 children for invitation around the age of 13 (early adolescence period).

The general design, all research aims and the specific measurements in the Generation R Study have been approved by the Medical Ethical Committee of the Erasmus Medical Center, Rotterdam. New measurements will only be embedded in the study after approval of the Medical Ethical Committee. Participants are asked for their written informed consent for the four consecutive phases of the study (prenatally, birth to 4 years, 4–16 years, and from 16 years onwards). At the start of each phase, mothers and their partners received written and oral information about the study. Even with consent of the parents, when the child is not willing to participate actively, no measurements are performed.

More information about the cohort can be found on the study website: https://generationr.nl

#### The Avon Longitudinal Study of Parents and Children

Pregnant women resident in Avon, UK with expected dates of delivery between 1st April 1991 and 31st December 1992 were invited to take part in the study.[4,5] A total number of 20,248 pregnancies have been identified as being eligible and the initial number of pregnancies enrolled was 14,541. Of the initial pregnancies, there was a total of 14,676 foetuses, resulting in 14,062 live births and 13,988 children who were alive at 1 year of age. The total sample size for analyses using any data collected after the age of seven is therefore 15,447 pregnancies, resulting in 15,658 foetuses. Of these 14,901 children were alive at 1 year of age.

Ethical approval for the study was obtained from the ALSPAC Ethics and Law Committee and the Local Research Ethics Committees. Informed consent for the use of all data collected was obtained from participants following the recommendations of the ALSPAC Ethics and Law Committee at the time. Participants can contact the study team at any time to retrospectively withdraw consent for their data to be used. Study participation is voluntary and during all data collection sweeps, information was provided on the intended use of data.

The completion of a questionnaire, either on paper or online, was considered to be written consent from participants to use their data for research purposes. For the majority of tests undertaken during face to face visits, verbal consent was obtained from participants (both parents and children as appropriate) prior to the start of any data collection. However, some tests required the completion of a written consent form. Biological samples are collected in accordance with the Human Tissue Act (2004). Specific Research Ethics Committee approval is sought for the consenting process at each collection sweep. Written consent, including permission for future use, is obtained from adult participants or from the parents of children as appropriate. Ethical approval for future use is covered by ALSPAC's Research Tissue Bank approval. All historical consents to hold biological samples have been reviewed as part of the Tissue Bank approval process. Participants can contact the study team at any time to retrospectively withdraw consent for use of their samples.

Please note that the study website contains details of all the data that is available through a fully searchable data dictionary and variable search tool: http://www.bristol.ac.uk/alspac/researchers/our-data/

**Flow chart of study participants**

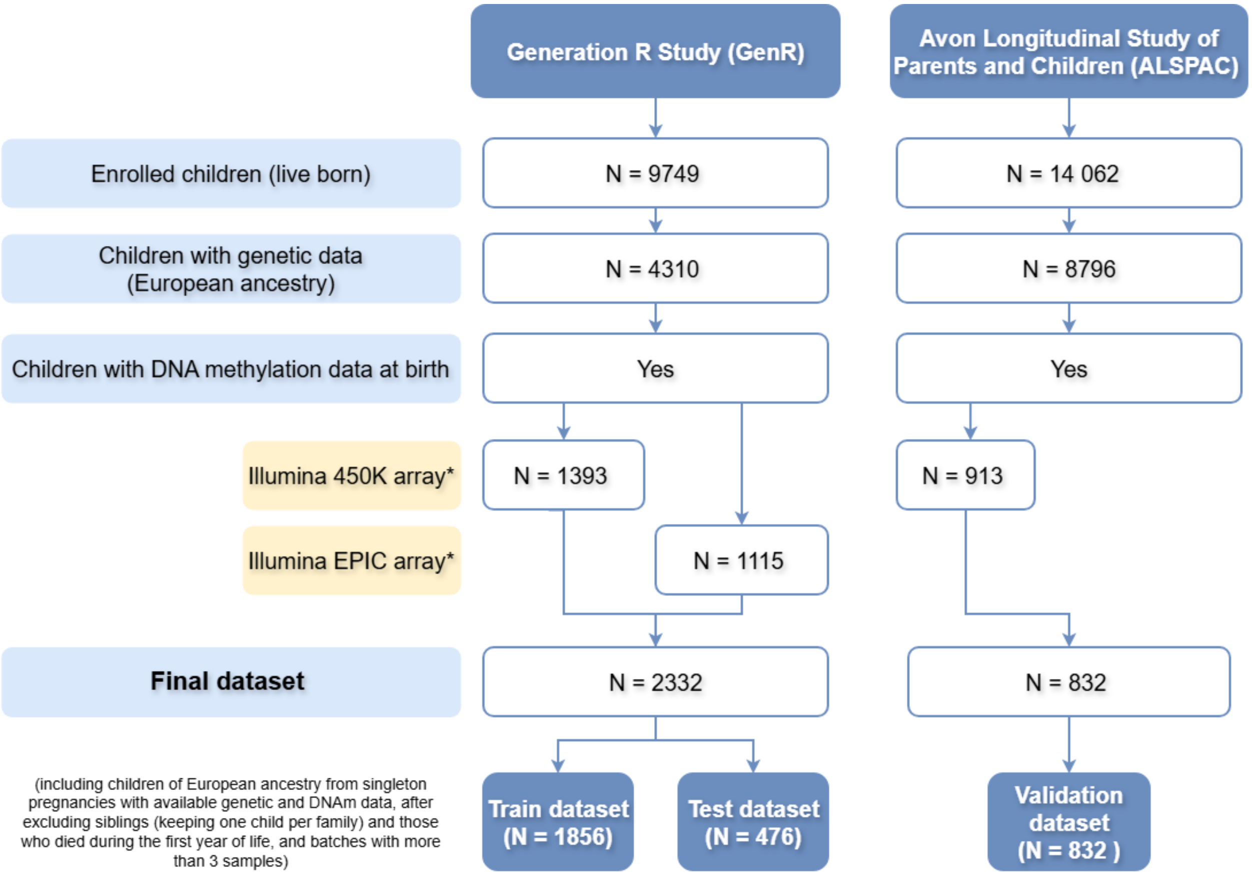

### Selection of prenatal risk factors

Literature search was conducted to identify prenatal risk factors that (1) have demonstrated associations with DNA, indicated by at least one CpG or DMR with p-value <0.05 adjusted for multiple comparisons (Bonferroni or FDR) in an epigenome-wide meta-analysis in cord blood or newborn blood and (2) have been shown to be associated with at least two of the three neurodevelopmental conditions (ADHD, autism and SCZ). The search strategy to identify articles to satisfy each of the two criteria was as follows.

| Criteria | PubMed Search strategy | Date of last update and number of articles found |
|---|---|---|
| 1.Epigenome-wide meta- analysis | (("DNA Methylation"[MeSH Terms] OR "methylation"[Title/Abstract] OR "epigenome-wide"[Title/Abstract])<br><br>AND<br><br>("perinatal"[Title/Abstract] OR "gestational"[Title/Abstract] OR "antenatal"[Title/Abstract] OR "pregnancy"[Title/Abstract] OR "birth"[Title/Abstract] OR "delivery"[Title/Abstract] OR "cord blood"[Title/Abstract] OR "newborn blood"[Title/Abstract])<br><br>Filters: in the last 10 years, Meta-Analysis, English, Humans, Child: birth-18 years | Date: 20.01.2026<br><br>Number of articles: 48<br><br>Additional articles identified through manual search: 1<br><br>Additional article on genetic liability for ADHD, autism and schizophrenia and DNAm: 1<br><br>Total articles: 50 |
| 2. Umbrella review or meta-analysis on prenatal and perinatal risk factors for ADHD, autism and schizophrenia | (Pregnancy[MeSH] OR pregnancy[Title/Abstract] OR prenatal[Title/Abstract] OR perinatal[Title/Abstract] OR gestation[Title/Abstract] )<br><br>AND<br><br>(Autism Spectrum Disorder[MeSH] OR Attention Deficit Hyperactivity Disorder[MeSH] OR Schizophrenia[MeSH] OR autism[Title/Abstract] OR ADHD[Title/Abstract] OR schizophrenia[Title/Abstract] )<br><br>AND<br><br>(environmental exposure*[Title/Abstract] OR environmental factor*[Title/Abstract] OR risk factor*[Title/Abstract] OR biomarker*[Title/Abstract] )<br><br>AND<br><br>(umbrella review[Title/Abstract] OR meta-analysis[Publication Type] OR meta-analysis[Title/Abstract] )<br><br>Filters: in the last 10 years, Meta-Analysis, Systematic Review, English, Humans | Date: 20.01.2026<br><br>Number of articles: 60 |

### Genotyping and calculation of polygenic scores (PGSs)

#### The Generation R Study

The Generation R Study Children's blood samples were collected from the umbilical cord at birth, or by venipuncture at around age 6 if cord blood was not available. Samples were preserved at −80 °C (umbilical) or −20 °C (venipuncture) and DNA was extracted using the Qiagen FlexiGene Kit. Detailed description of the cohort's biological sample collection procedures, sample processing and storage and available measurements have been published previously.[6,7] Participants of the Generation R Study were genotyped in two rounds, creating two subsets: GENR3 and GENR4. This study used only the GENR3 subset, as it was the one with both genetic data and DNA methylation data at birth.

GENR3 was genotyped using Illumina HumanHap 610 or 660 Quad chips. The two Genome Studio projects (one each for the HumanHap 610 array and for the 660 array), were merged using SNPs common to both arrays. The quality control (QC) procedure included marker QC and sample QC. Marker QC filtering included (1) marker call rate 95%, (2) minor allele frequency (MAF) ≤ 0.001 (3) differential missingness between the two projects, (4) deviation from Hardy–Weinberg equilibrium proportion ($P < 10^{-7}$). Sample QC included (1) duplicate detection, (2) sex discordance (3) call rate 97.5% and (4) high heterozygosity. After QC, the genotyped markers were phased using MACH and then imputed using the Minimac software to the 1000 Genomes Project Phase III version 5 reference.

The GENR3 subset of genome-wide-variants used to calculate the PGS included 7673245 SNPs, after filtering for imputation quality > 0.8 and MAF > 1% and excluding multi-allelic SNPs, duplicate variants and structural variants (indels). The first 20 genomic principal components were calculated for both European or non-European GENR3 children. More detailed description of the quality control, phasing, imputation and analytical considerations for GENR3 have been described before.[8]

#### The Avon Longitudinal Study of Parents and Children

Genetic data for ALSPAC children were derived using the Illumina HumanHap550 quad chip from cord blood. Genotype calls were made using Illumina GenomeStudio software. The quality control (QC) procedure included marker QC and sample QC. Individuals were excluded on the basis of (1) gender mismatches, (2) minimal or excessive heterozygosity, (3) disproportionate levels of individual missingness (>3%) and (4) insufficient sample replication (IBD < 0.8). SNPs QC included with a (1) MAF < 1%, (2) call rate of < 95% or evidence for violations of Hardy-Weinberg equilibrium ($P < 10^{-7}$). After combining the genotypes of mothers and children, SNPs with genotype missingness above 1% were removed and subjects due to potential ID mismatches

Haplotypes were estimated using ShapeIT(v2.r644) which utilizes relatedness during phasing. Phased version of the 1000 genomes reference panel was obtained (Phase 1, Version3) from the Impute2 reference data repository (phased using ShapeItv2.r644, haplotype release date Dec 2013). Imputation of the target data was performed using Impute V2.2.2 against the reference panel (all polymorphic SNPs excluding singletons), using all 2186 reference haplotypes (including non-Europeans).

The ALSPAC subset of genome wide-variants used to calculate the PGS included 7355830 SNPs, after filtering for imputation quality > 0.8 and MAF > 1% and excluding multi-allelic SNPs, duplicate variants, structural variants (indels) and 199 SNPs reported to have incorrect strand annotations in the December 2013 release of 1000 Genomes phase 1 version 3. The first 20 genomic principal components were calculated. More detailed description of the quality control, phasing, imputation and analytical considerations for ALSPAC are available at: ALSPAC OMICs Data Catalogue

#### Calculation of PGSs

In both Generation R and ALSPAC, the PGSs for ADHD, autism and schizophrenia were calculated using a clumping and thresholding method PRSice-2.[9] The default options were used, meaning that the correlated SNPs were clumped within a 250 kb window at a R2 threshold of 0.1. PRSice2 allows the PGSs to be calculated at multiple thresholds. The optimal PGS thresholds were identified in a previous study[10] that, among other cohorts, included Generation R and ALSPAC. The optimal threshold for ADHD and autism was chosen by meta-analyzing associations between the PGSs at multiple thresholds and their corresponding diagnosis-related measures. The thresholds chosen were 0.01 for ADHD (meta-analysis between PGS and phenotype $p=9.8x10^{-34}$) and 0.5 for autism (meta-analysis of association between PGS and phenotype $p=1.2x10^{-6}$). Due to the lack of diagnosis-related measures at an earlier age for schizophrenia, instead of performing parameter optimization the study used preselected threshold of 0.05 in accordance with prior research.

### Prenatal risk factors

#### Generation R Study

**Maternal age** was self-reported and collected from questionnaires sent out during pregnancy. **Maternal pre-pregnancy BMI** was calculated by using the weight in kilograms (kg) divided by height in meters (m) squared. Pre-pregnancy maternal weight was self-reported, while maternal height (cm) and weight (kg) were measured at enrolment. BMI calculated using self-reported weight and measured height was used when available. Otherwise, when mothers enrolled before 13th gestational week, BMI was calculated using measured weight and height at enrollment. The correlation between the two BMI measures is 0.956. **Maternal smoking during pregnancy** was assessed using questionnaires at enrolment. Mother who were enrolled before a gestational age <18 and between 18 and 25 weeks of gestation, also received a second and third trimester questionnaire, and third trimester only questionnaire, respectively. Using data from all questionnaires, smoking was categorized into never smoked in pregnancy, smoking until pregnancy was known, and continued smoking in pregnancy. In this study, we used the variable "sustained maternal smoking" categorized as (1) never smoked during pregnancy or smoked until pregnancy was known and (2) continued smoking in pregnancy. **Gestational diabetes** was diagnosed by a community midwife or an obstetrician according to Dutch midwifery and obstetric guidelines. The presence of gestational diabetes was retrieved from birth records after delivery. In the Netherlands it is advised that, in case of gestational diabetes, antenatal care and delivery takes place under the responsibility of an obstetrician. Information on **Hypertensive Disorders of Pregnancy** was obtained in multiple ways. Information about gestational hypertension abstracted from the medical records and derived according to the International Society for the Study of Hypertension in Pregnancy (ISSHP) criteria. According to the Dutch standards of antenatal care, all women whose pregnancies are complicated by preeclampsia should deliver in a hospital under medical supervision of an obstetrician. The delivery reports of study participants who delivered under medical supervision were retrieved and screened by a trained medical record abstractor. To confirm the presence of preeclampsia, the same abstractor conducted detailed reviews of hospital charts of these women. Preeclampsia was defined according to the criteria described by the ISSHP. **Gestational age at birth** was obtained as follows. Participating pregnant mothers in Generation R Study were seen in the first trimester of pregnancy for fetal ultrasound at our research center. During this visit, gestational age was established. If mothers had a known and reliable first day of the LMP, and a regular menstrual cycle of 28 ± 4 days, the clinical estimate of gestational age was based on LMP. If mothers did not know the exact date of their LMP, or had an irregular menstrual cycle, we established gestational age by ultrasound examination, which does not take into account variation in early fetal growth. For women who did not attend the prenatal visits, gestational age was retrieved from the Netherlands National Obstetric Register. **Birthweight** was obtained from midwife and obstetric records. **Mode of delivery** was obtained from midwife and obstetric records. The definition of Cesarean Delivery includes both planned and unplanned cesarean delivery.

##### Avon Longitudinal Study of Parents and Children

**Maternal age** was self-reported and collected from questionnaires sent out during pregnancy. **Maternal pre-pregnancy BMI** was calculated by using the weight in kilograms (kg) divided by height in meters (m) squared. Pre-pregnancy weight and height was obtained through a questionnaire. **Maternal smoking during pregnancy** was assessed through questionnaires administered at 18 and 32 weeks of gestation. From these data, smoking in pregnancy was classified as no smoking, smoking in first trimester only and continued smoking. In this study, we used the variable "sustained maternal smoking" categorized as (1) never smoked during pregnancy or only in first trimester and (2) continued smoking in pregnancy (any smoking during pregnancy, except for smoking only in first trimester). Physician diagnosed **gestational diabetes** was obtained through obstetric records. **Gestational age at birth** was retrieved from the obstetric records. If there was any suggestion (based on last menstrual period or ultrasound measures) that the gestation was preterm, clinical records were reviewed again. Preterm birth was defined as delivery occurring at less than 37 weeks of gestation. **Birthweight** was obtained from midwife and obstetric records. **Mode of delivery** was obtained from midwife and obstetric records. The definition of cesarean delivery includes both planned and unplanned cesarean delivery.

#### Childhood psychiatric outcomes

##### The Generation R Study

1. ***Repeated measures of broadly defined childhood psychiatric problems***

The **Child Behaviour Checklist (CBCL)** was used to ascertain parent-reported emotional and behavioural problems during the past six months. In this study, CBCL administered and completed by the primary caregiver at approximate **ages of 6, 10 and 14** were used. Two age-appropriate versions of the CBCL were used: the preschool version (CBCL/1.5-5) (Achenbach & Rescorla, 2000) up to the age of 6 and the school-age version (CBCL/6-18) (Achenbach & Rescorla, 2001) for the ages after that. For the wave at 6 years, the CBCL/1.5-5 was used, as 58% of the children were 5 years old at the assessment round. For both the preschool and school version, **Total Problems, Internalising Problems, Externalising Problems scales and Attention Problems** were calculated. Total Problems is calculated by summing all individual items. In the preschool version, Internalising Problems scores were derived from the Anxious/Depressed, Withdrawn, Somatic Complaints, and Emotionally Reactive scales, whereas in the school-age version, they were based on Anxious/Depressed, Withdrawn/Depressed, and Somatic Complaints scales. Externalising Problems were derived by summing up items from the Aggressive Behaviour and Attention Problems scales in the preschool version, and the Rule-Breaking Behaviour and Aggressive Behaviour scales in the school-age version. For each scale, sum scores were transformed into t-scores.

2. ***Autistic traits***

The **Social Responsiveness Scale** (SRS) was used to measure autistic traits in children over the past six months (Constantino & Gruber, 2005; Constantino et al., 2000) at aproximate **age of 6**. To minimize subject burden, we assessed an 18-item short version of the scale. (Román et al., 2013) The SRS consists of three subscales, assessing Social Cognition (5 items), Social Communication (8 items), and Mannerism (5 items). The abbreviated version of the SRS has been shown to correlate highly (correlation = .99) with the full SRS version (Román et al., 2013).

##### 3. *Attention-Deficit/Hyperactivity Disorder (ADHD) traits*

The **Conners' Parent Rating Scale–Revised Short Form** (Conners et al., 1998) was used to assess ADHD traits in children at approximate **age of 8**. This questionnaire contains 27-items assessing attention problems and symptoms of oppositional defiant disorder. This version of the Conners' Parent Rating Scale includes 4 scales: Cognitive Problems/Inattention (6 items), Hyperactivity (6 items), Oppositional (6 items), and the ADHD index (ADHDi; 12 items, which also includes 3 items of the Cognitive Problems/Inattention scale). The ADHD index was used to assess ADHD traits.

##### 4. *Delusions*

Lifetime experience of delusions was self-reported at age 14, using 6 items relating specifically to delusional experiences from the Kiddie-Schedule for Affective Disorders and Schizophrenia (K-SADS; Kaufman et al., 1997; Townsend et al., 2020) which was adapted for use in a self-reported questionnaire (Adriaanse et al., 2015) and included beliefs that one's thoughts could be read, ideas of reference, beliefs about being spied on, under control of special power, having grandiose abilities and delusion of bodily change. Items were rated on a 3-point Likert scale (1 = no, 2 = yes, likely, 3 = yes, definitely). The total delusions score included the sum of all item scores.

##### 5. *Hallucinations*

Hallucinatory experiences from the last six months were self-reported at age 14 using two items from the Youth Self-Report (YSR; Achenbach, 2001). The specific items were "I hear sounds or voices that other people think aren't there" and "I see things that other people think aren't there". The sum score was dichotomized into a binary variable of any hallucinatory experiences (Steenkamp et al., 2021).

#### The Avon Longitudinal Study of Parents and Children

##### 1. *Repeated measures of broadly defined childhood psychiatric problems*

The parent-reported **Strengths and Difficulties Questionnaire** (SDQ) measures children's psychosocial development in five domains: emotional problems, peer problems, conduct problems, hyperactivity/inattention, and prosocial behavior. In this study, the SDQ administered and completed by the primary caregiver in ALSPAC at the approximate **ages of 7, 10 and 13** was used. **Total Problems, Internalising Problems, Externalising Problems scales and Hyperactivity/ Inattention Problems** were calculated. Total Problems scores were calculated using the prorated sum of the hyperactivity/inattention, emotional symptoms, conduct problems, prosocial behavior, and peer problems subscales. Internalizing Problems were derived using the prorated sum of the emotional problems and peer problems subscales, while Externalizing Problems were derived using the prorated sum of the conduct problems and hyperactivity/inattention problems.

##### 2. *Autistic traits*

Autistic traits were measured using the parent-rated 10-item **Social Communication Disorder Checklist (SCDC)**, completed at **age 7**. The questionnaire is designed to assess social reciprocity as well as verbal and nonverbal communication. The questionnaire score ranges from 0 to 24 and has high specificity and sensitivity for autism diagnosis.

3. ***Attention-Deficit/Hyperactivity Disorder (ADHD) traits***

ADHD traits were assessed at **age 7** using the parental report from the **Development and Well-Being Assessment (DAWBA)**. The DAWBA is a validated instrument that combines structured and semi-structured questions aligned with DSM-IV and ICD diagnostic criteria. DAWBA has shown good predictive value for ADHD. We used the DAWBA DSM-IV 6-band computer prediction for "any ADHD disorder"

4. ***Delusions and Hallucinations***

Delusions and hallucinations were assessed using the **psychosis-like experience (PLIKS)** semi-structured interview when they were **13 years of age**. The interview includes 12 core questions assessing the occurrence of hallucinations and psychotic experiences in the past six months. The questionnaire has two questions covering visual and auditory hallucinations, and dichotomized into a binary variable of any hallucinatory experiences. Delusions included beliefs that one's thoughts could be read, ideas of reference, beliefs about being spied on, under control of special power and having grandiose abilities. Interviewers rated symptom as either not present, suspected or definitely present, and the total delusions score was the sum of items covering delusions.

### Covariates

#### The Generation R Study

The **sex of the child** was determined at birth by attending midwives. **Maternal education** (as proxy for socio-economic status) was assessed by a questionnaire and defined by the highest attained educational level obtained and was dichotomized into high or low educational attainment. High educational attainment was defined as obtaining Bachelor's degree or higher academic education. **Ancestry** was accounted for by incorporating the first principal components derived from the genotype data. **Cell type proportions**, namely CD8 T cells, CD4 T cells, natural killer cells, B cells, monocytes, granulocytes and nucleated red blood cells, were estimated from DNA methylation values, were estimated using the Gervin method[12]. **Batch effects** were accounted for by including the sample plate as a covariate.

#### The Avon Longitudinal Study of Parents and Children

The **sex of the child** was determined at birth by attending midwives. **Maternal education** (as proxy for socio-economic status) was assessed by a questionnaire and defined by the highest attained educational level obtained and was dichotomized into high or low educational attainment. High educational attainment was defined as obtaining a university degree or A-level education. **Ancestry** was accounted for by incorporating the first principal components derived from the genotype data. **Cell type proportions**, namely CD8 T cells, CD4 T cells, natural killer cells, B cells, monocytes, granulocytes and nucleated red blood cells, were estimated from DNA methylation values, were estimated using the Gervin method.[12] **Batch effects** were accounted for by including the sample plate as a covariate.

### DNA methylation measurement

#### The Generation R Study

DNA extracted (using the salting-out method) from blood samples taken at birth (cord blood) was used for this analysis. 500 ng DNA per sample underwent bisulfite conversion using the EZ-96 DNA Methylation kit (Shallow) (Zymo Research Corporation, Irvine, USA). Samples were plated onto 96-well plates in no specific order. Samples from the first bwere processed with the e Illumina Infinium HumanMethylation450 BeadChip (Illumina Inc., San Diego, USA) which analyses methylation at >450 000 CpG sites (GENR-450K) or Illumina Infinium MethylationEPIC v1.0 BeadChip (Illumina Inc., San Diego, USA) which analyses methylation at >850 000 CpG sites (GENR-EPIC), giving rise to two distinct subsets of epigenetic data that were processed independently, using the same workflow, as described below.

Preparation and normalization of the data was performed according to the CPACOR workflow[14] using the software package R[15]. In detail, the idat files were read using the minfi package[16]. Probes that had a detection p-value above background (based on sum of methylated and unmethylated intensity values) ≥ 1E-16 were set to missing per array. Next, the intensity values were stratified by autosomal and non-autosomal probes and quantile normalized for each of the six probe type categories separately: type II red/green, type I methylated red/green and type I unmethylated red/green. Beta values were calculated as proportion of methylated intensity value on the sum of methylated+unmethylated+100 intensities. Arrays with observed technical problems such as failed bisulfite conversion, hybridization or extension, as well as arrays with a mismatch between sex of the proband and sex determined by the chr X and Y probe intensities were removed from subsequent analyses. Additionally, only arrays with a call rate > 96% per sample were processed further. Probes on the X and Y chromosomes were excluded from the dataset.

The final GENR-450K dataset contains information on 458,563 CpGs for 1393 samples at birth. The final GENR-EPIC dataset contains information on 808,183 CpGs for 1115 samples at birth.

#### The Avon Longitudinal Study of Parents and Children

Our study population arises from the Accessible Resource for Integrated Epigenomics Studies (ARIES) project,[17] a sub-study drawn from the ALSPAC mother-child cohort.[4,5]

DNA methylation was quantified using the Illumina Infinium HumanMethylation450K BeadChip assay (ALSPAC-450K) (Illumina Inc., CA). Cord blood (collected immediately after birth) and peripheral blood samples (whole blood or buffy coat) were collected according to standard procedures. Following extraction, DNA was bisulfite-converted using the Zymo EZ DNA MethylationTM kit (Zymo, Irvine, CA) then genome-wide methylation status of over 485 000 CpG sites was measured using the Illumina 450 K array according to the standard protocol.

Samples from all participant ages in ARIES were distributed across slides using a semi-random approach (sampling criteria were in place to ensure that all time points were represented on each array) to minimize the possibility of confounding by batch effects. The QC process was conducted using the meffil package.[18] QC steps included checks for genotype and sex mismatches, incorrect relatedness, sample concordance at different time points, dye bias, and probe detection efficiency. The Illumina 450 K BeadChip assay detects the proportion of molecules methylated at each CpG site on the array. For each sample, the estimated methylation level at each CpG site was expressed as a beta value (β), which is the ratio of the methylated probe intensity and the overall intensity and ranges from 0 (no cytosine methylation) to 1 (complete cytosine methylation).

Methylation data were pre-processed using the R software[15] and betas were normalized using functional normalization implemented in the meffil package.[18] Ten 10 principal compomnents on the controlmatrix were used to regress out technical variation. Slide was regressed out as random effect before normaliziation.

12. Gervin K, Salas LA, Bakulski KM, et al. Systematic evaluation and validation of reference and

library selection methods for deconvolution of cord blood DNA methylation data. *Clin Epigenetics*. 2019;11(1):125. doi:10.1186/s13148-019-0717-y
