## Supplementary Figure for "DNA methylation as proxy of genetic, prenatal and perinatal psychiatric risk factors"

**Supplementary Figure 1.** Distribution of continuous risk factors across datasets.

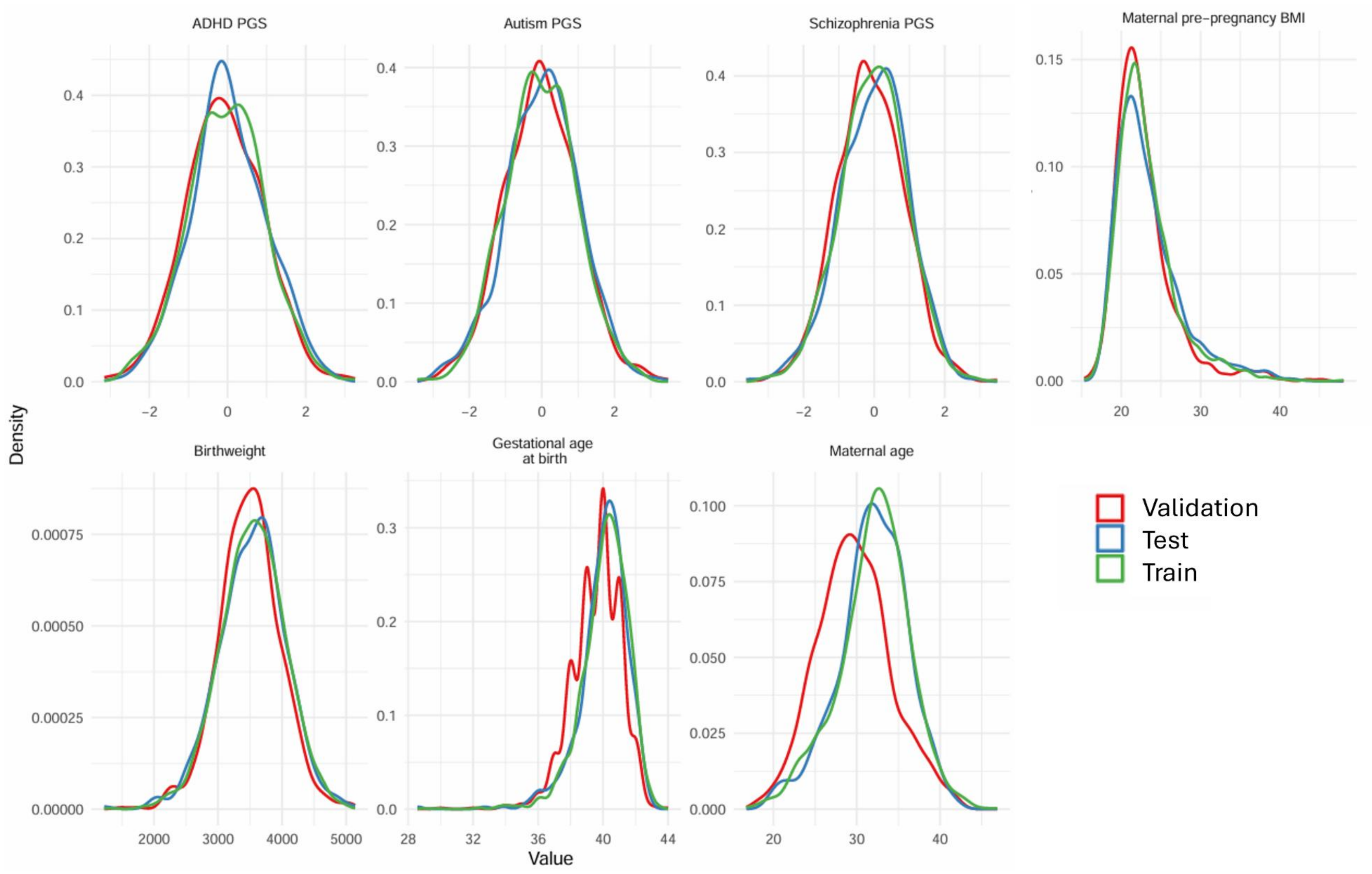

**Supplementary Figure 2.** Distribution of binary risk factors (and hallucinations) across datasets

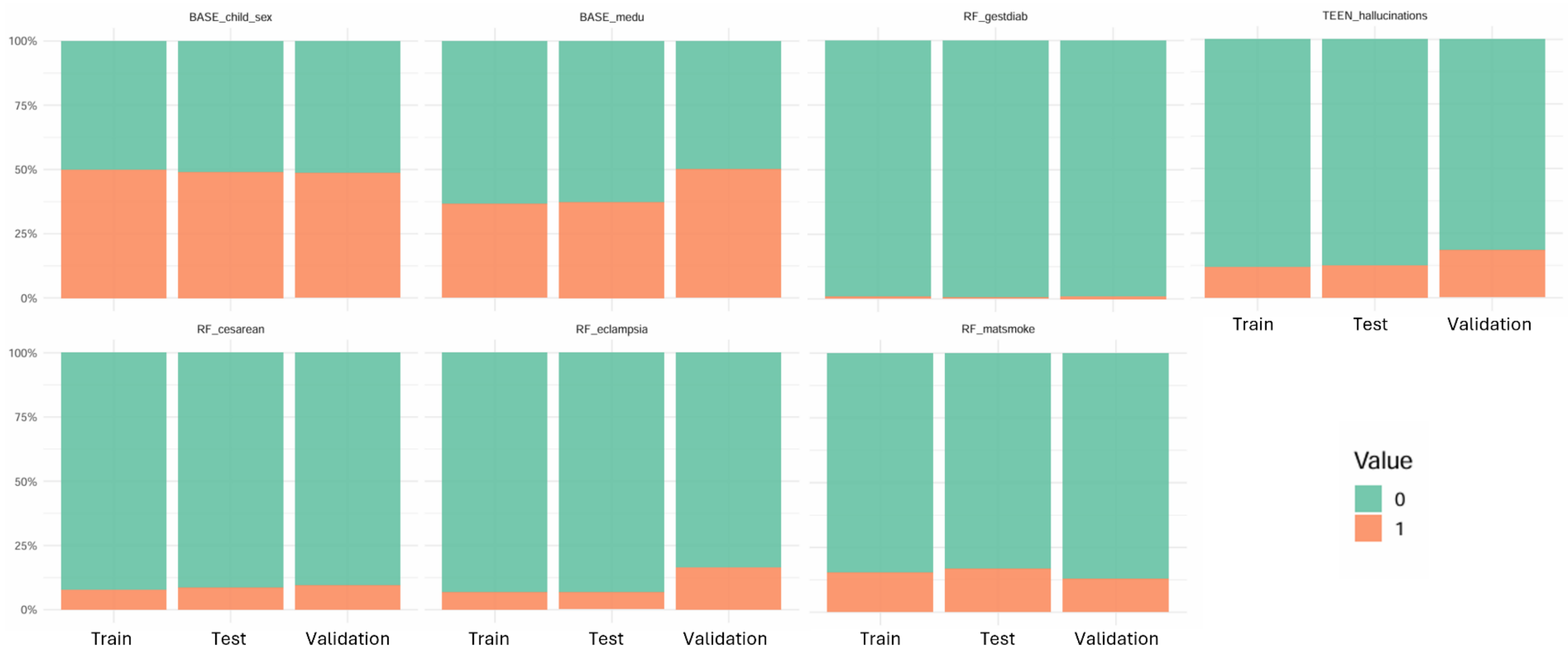

**Supplementary Figure 3.** Distribution of outcomes across datasets. Continuous outcomes were standardized as z-scores to enable comparability, given that Generation R (training/test) and ALSPAC (validation) use different instruments to measure.

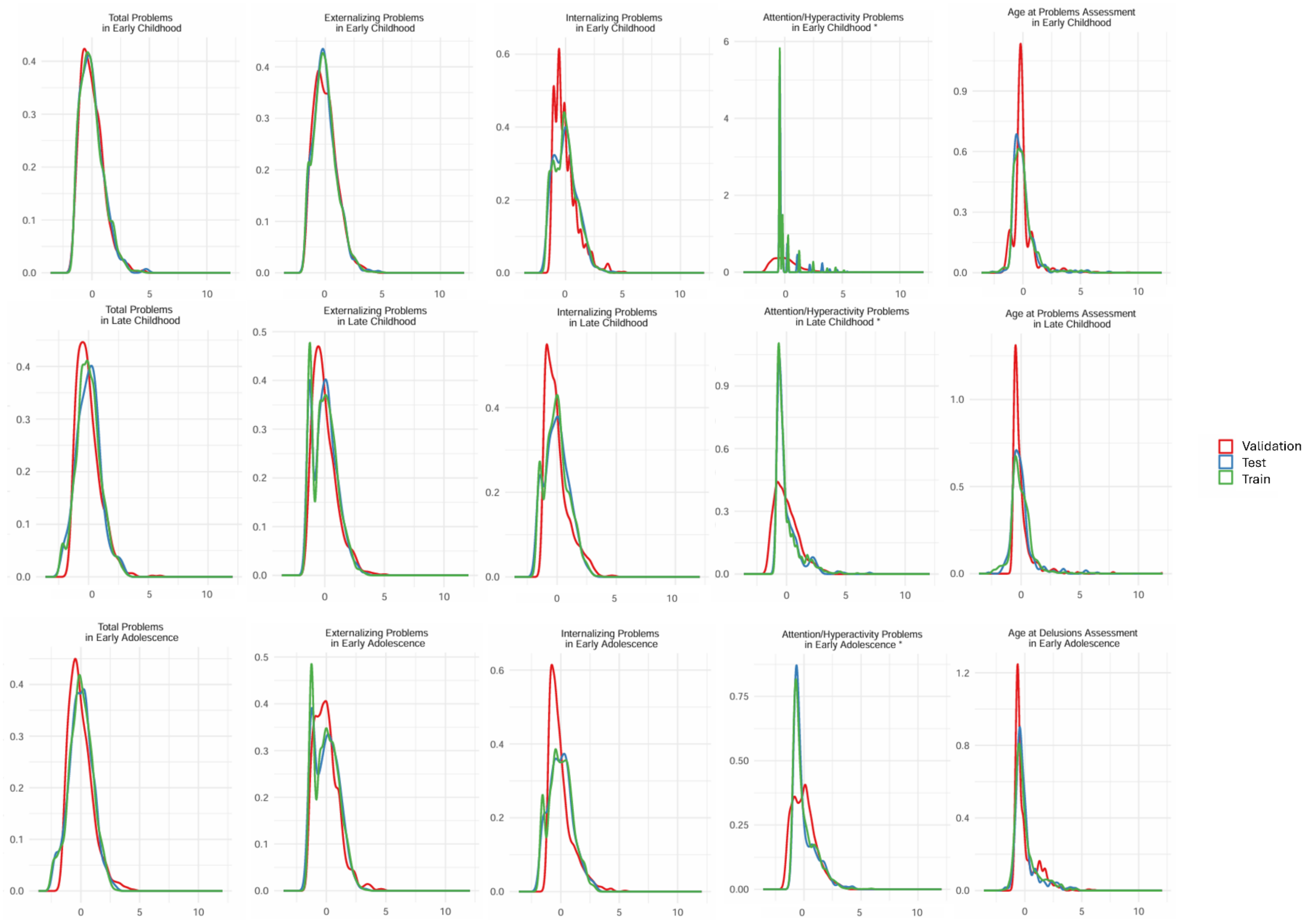

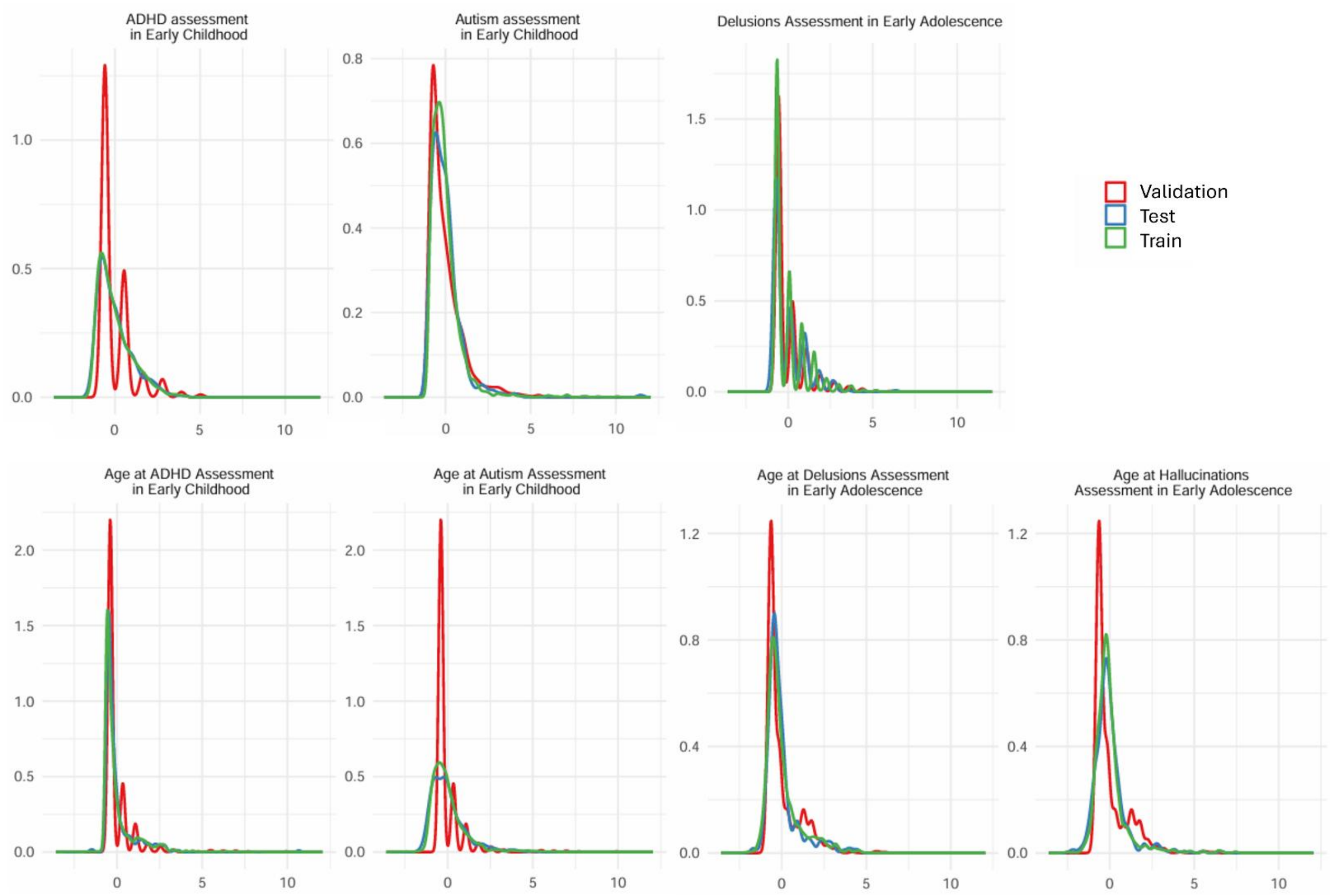
ADHD assessment
in Early Childhood
Autism assessment
in Early Childhood
Delusions Assessment in Early Adolescence
Validation
Test
Train
Age at ADHD Assessment
in Early Childhood
Age at Autism Assessment
in Early Childhood
Age at Delusions Assessment
in Early Adolescence
Age at Hallucinations
Assessment in Early Adolescence

**Supplementary Figure 4.** Percentage of missing values across main variables across datasets.

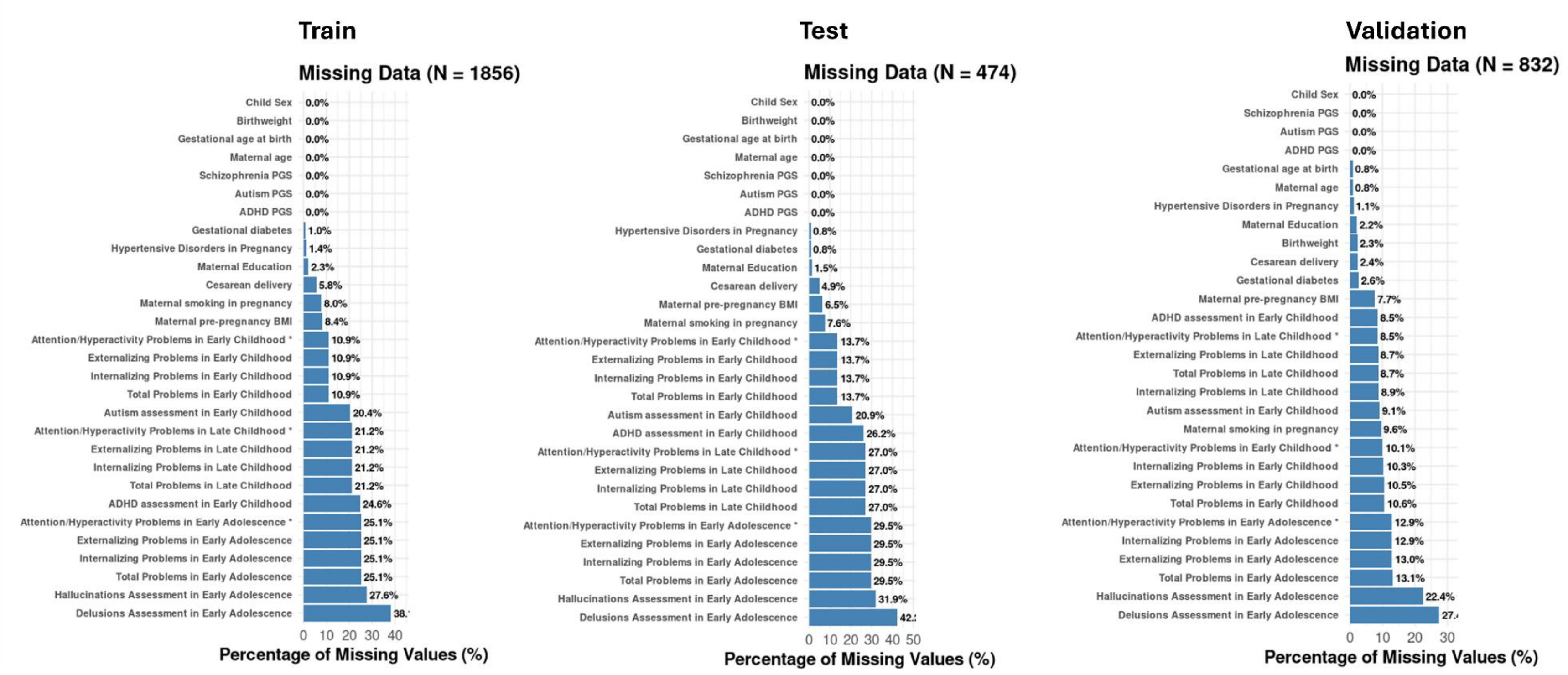

**Supplementary Figure 5. Correlations** between genetic and prenatal risk factors and their corresponding risk factor methylation profile scores (RF-MPSs) in the **original (non-imputed) dataset**. The displayed correlations indicate p-value<0.05.

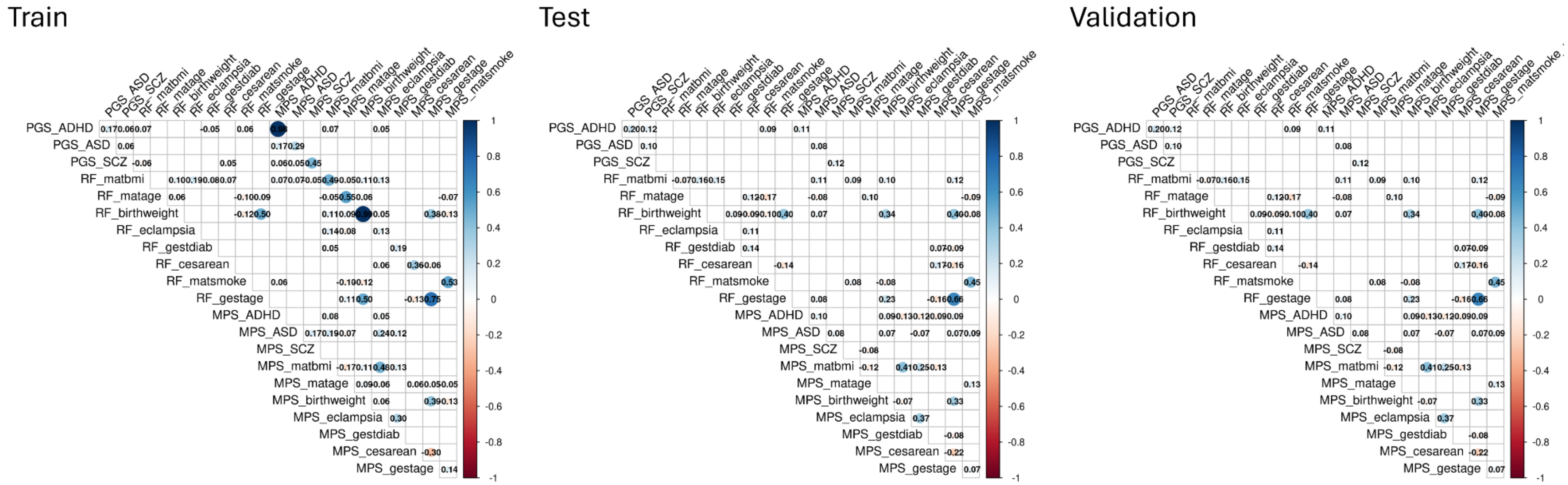

**Supplementary Figure 6. Covariate-adjusted partial correlations** between genetic and prenatal risk factors and their corresponding risk factor methylation profile scores (RF-MPSs) in the **original (non-imputed) dataset**. The displayed correlations indicate p-value<0.05.

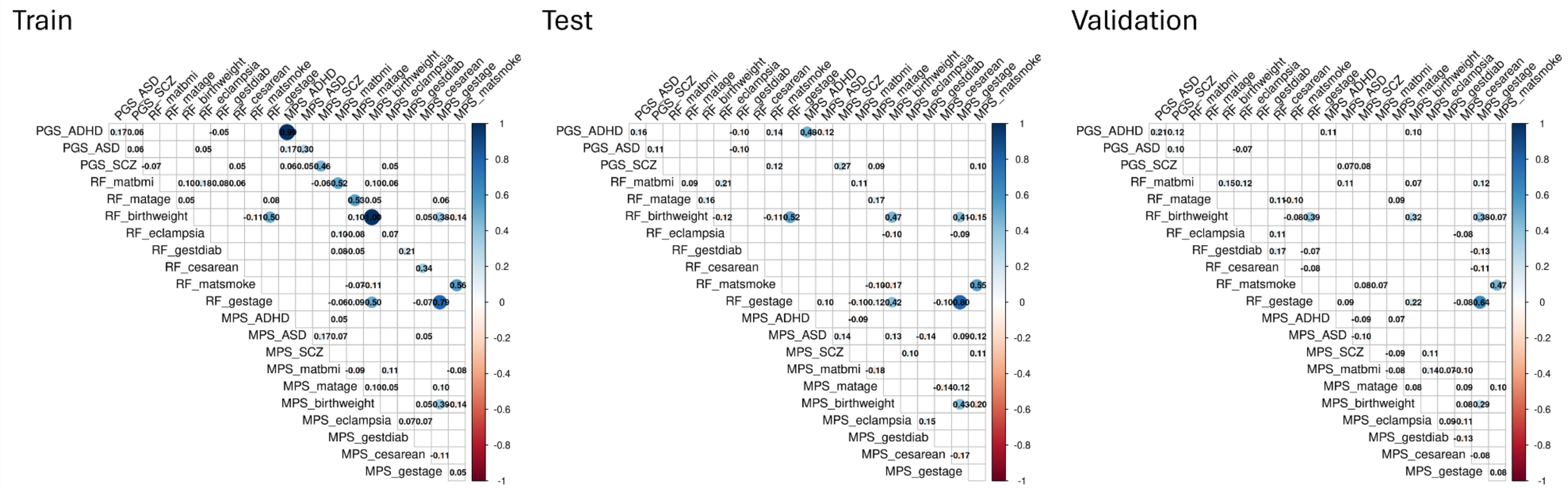

**Supplementary Figure 7. Correlations** between genetic and prenatal risk factors and their corresponding risk factor methylation profile scores (RF-MPSs) in the **imputed dataset**. The displayed correlations indicate p-value<0.05.

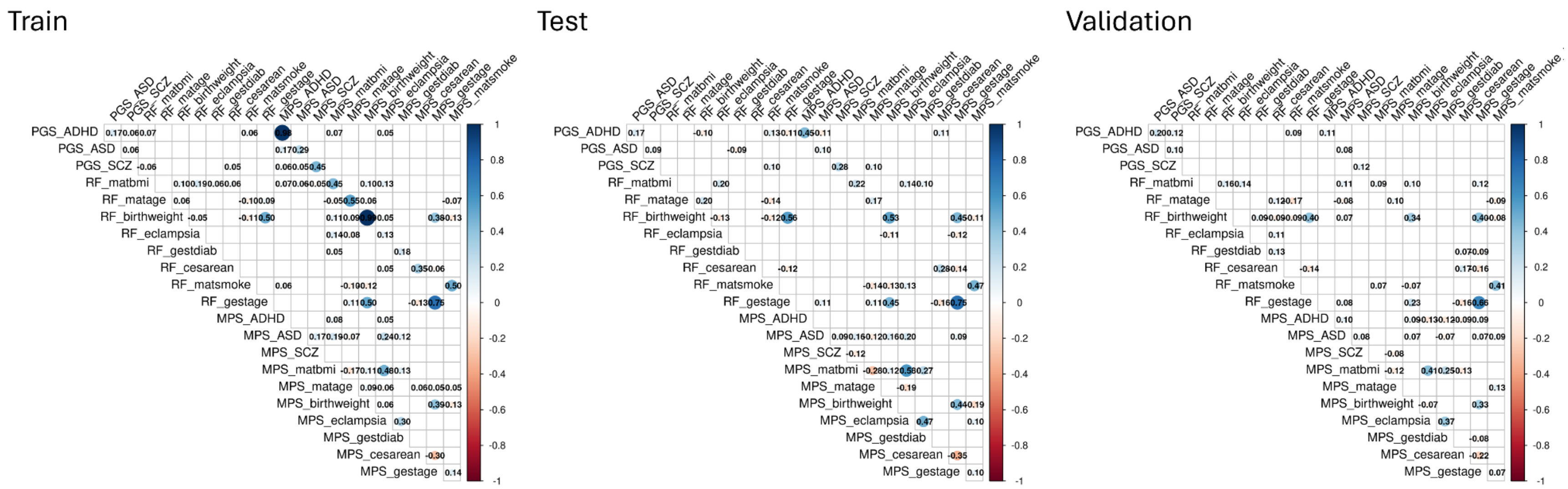

**Supplementary Figure 8. Covariate-adjusted partial correlations** between genetic and prenatal risk factors and their corresponding risk factor methylation profile scores (RF-MPSs) in the **imputed dataset**, also reported in the main table. The displayed correlations indicate p-value<0.05.

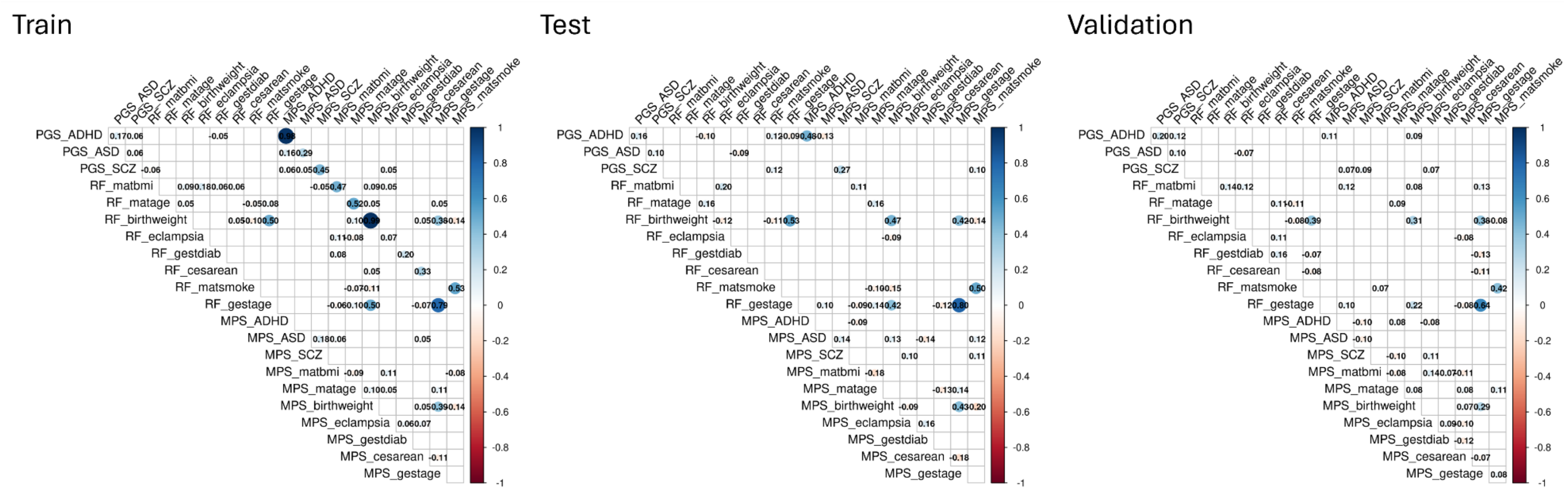
